# Effectiveness of mRNA and ChAdOx1 COVID-19 vaccines against symptomatic SARS-CoV-2 infection and severe outcomes with variants of concern in Ontario

**DOI:** 10.1101/2021.06.28.21259420

**Authors:** Sharifa Nasreen, Hannah Chung, Siyi He, Kevin A. Brown, Jonathan B. Gubbay, Sarah A. Buchan, Deshayne B. Fell, Peter C. Austin, Kevin L. Schwartz, Maria E. Sundaram, Andrew Calzavara, Branson Chen, Mina Tadrous, Kumanan Wilson, Sarah E. Wilson, Jeffrey C. Kwong, on behalf of the Canadian Immunization Research Network (CIRN) Provincial Collaborative Network (PCN) Investigators

## Abstract

SARS-CoV-2 variants of concern (VOC) are more transmissible and have the potential for increased disease severity and decreased vaccine effectiveness. We estimated the effectiveness of BNT162b2 (Pfizer-BioNTech Comirnaty), mRNA-1273 (Moderna Spikevax), and ChAdOx1 (AstraZeneca Vaxzevria) vaccines against symptomatic SARS-CoV-2 infection and COVID-19 hospitalization or death caused by the Alpha (B.1.1.7), Beta (B.1.351), Gamma (P.1), and Delta (B.1.617.2) VOCs in Ontario, Canada using a test-negative design study. Effectiveness against symptomatic infection ≥7 days after two doses was 89–92% against Alpha, 87% against Beta, 88% against Gamma, 82–89% against Beta/Gamma, and 87–95% against Delta across vaccine products. The corresponding estimates ≥14 days after one dose were lower. Effectiveness estimates against hospitalization or death were similar to, or higher than, against symptomatic infection. Effectiveness against symptomatic infection is generally lower for older adults (≥60 years) compared to younger adults (<60 years) for most of the VOC-vaccine combinations.

## Introduction

Evolution of the SARS-CoV-2 virus over the course of the COVID-19 pandemic has resulted in the emergence of four variants of concern (VOC), Alpha (B.1.1.7), Beta (B.1.351), Gamma (P.1), and Delta (B.1.617.2), to date. Alpha was first detected in United Kingdom (UK) in September 2020, Beta in South Africa in May 2020, Gamma in Brazil in November 2020, and Delta in India in October 2020^1^. As of 28 September 2021, 193, 142, 96, and 187 countries, territories, and areas globally have reported Alpha, Beta, Gamma, and Delta cases respectively^2^. SARS-CoV-2 variants of concern (VOC) are more transmissible and have the potential to cause increased disease severity and to decrease COVID-19 vaccine effectiveness^1^. As of the time of writing, Delta is now the dominant VOC that is driving increases in infections, even in populations with high levels of access to vaccines.

Evidence of the efficacy of COVID-19 vaccines against VOCs from randomized clinical trials is mostly limited to post-hoc estimates of vaccine efficacy against symptomatic infection; however, these analyses suffer from insufficient statistical power and may not have variant information for all cases. Vaccine efficacy against symptomatic infection >14 days after the second dose was reported to be 70% (95% confidence interval [CI]: 44 to 85%)^3^ against Alpha and 10% (95% CI, -77 to 55%)^4^ against Beta for ChAdOx1 (AstraZeneca Vaxzevria). Efficacy against infection by Beta >7 days after the second dose for BNT162b2 (Pfizer-BioNTech Comirnaty) was 100% (95% CI, 54 to 100%)^5^. Few observational studies have reported the effectiveness of COVID-19 vaccines against infection or severe outcomes caused by VOCs^6-11^.

Ontario, the largest province in Canada, implemented a three-phased COVID-19 vaccination program starting in December 2020 and adopted a delayed second-dose strategy due to vaccine supply constraints during phase 1^12^. BNT162b2 became available on 14 December 2020, mRNA-1273 (Moderna SpikeVax) on 28 December 2020, and ChAdOx1 on 10 March 2021^13^. All four VOCs have been circulating at various times in Ontario in 2021^14^. Our objective was to estimate the effectiveness of BNT162b2, mRNA-1273, and ChAdOx1 vaccines against symptomatic SARS-CoV-2 infection and severe outcomes (COVID-19 hospitalization or death) caused by Alpha, Beta, Gamma, and Delta between December 2020 and August 2021 in Ontario.

## Results

### Study population

Over the study period, we identified 682,071 symptomatic community-dwelling individuals who were tested for SARS-CoV-2, with 31,440 (5%) positive for non-VOC SARS-CoV-2 and 51,440 (8%) positive for a VOC (Table 1). Cases of Delta were younger, more likely to reside in Central West region, more likely to occur later in the study period, less likely to have any comorbidities, and more likely to reside in neighbourhoods with lower household income and greater proportions of essential workers than cases of other VOCs and non-VOC SARS-CoV-2 infections, as well as test-negative controls.

**Table 1:**
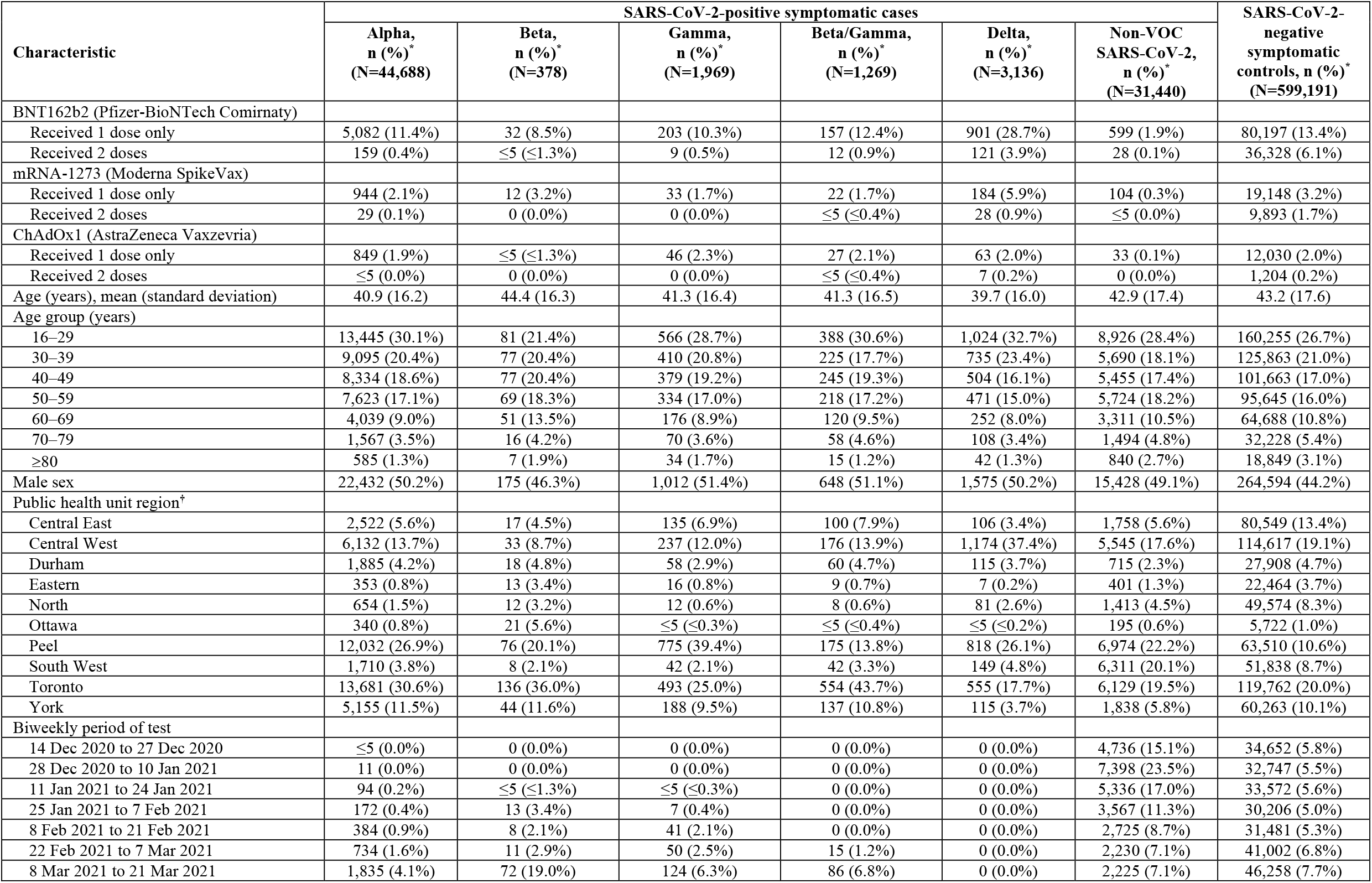
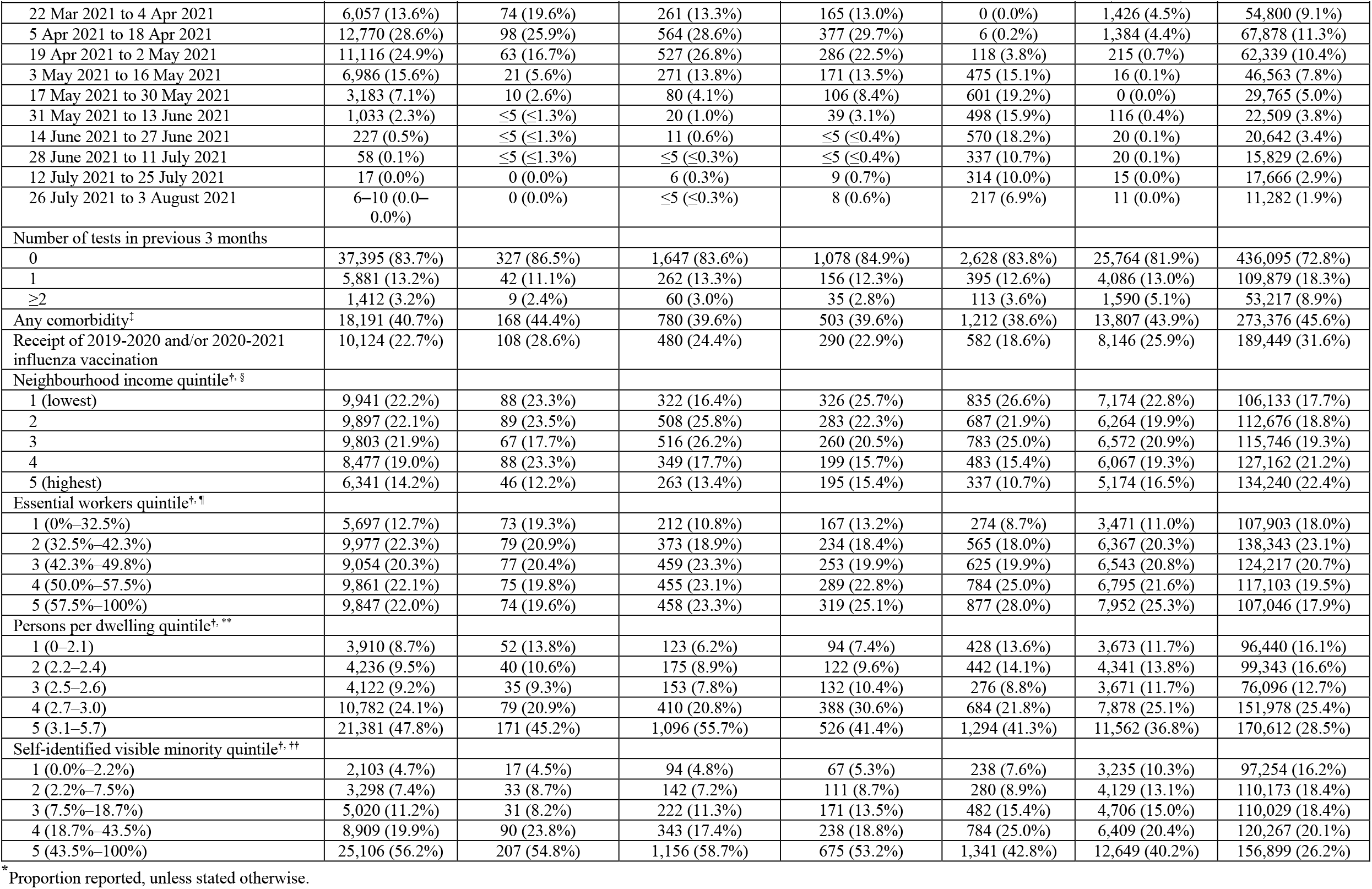
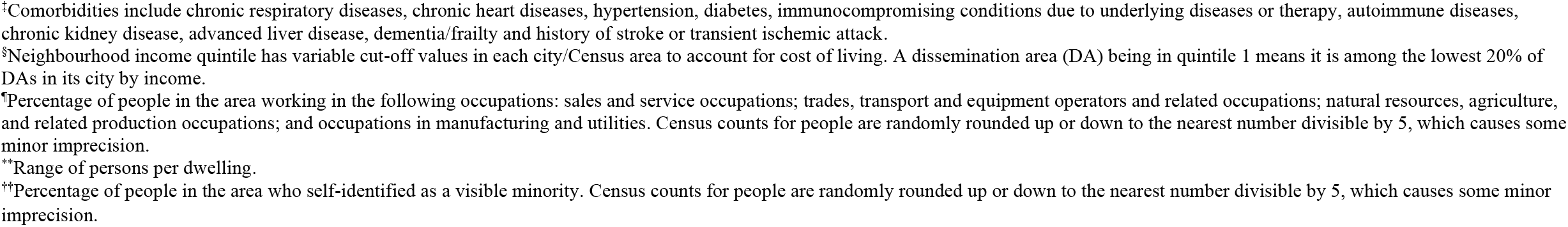
Characteristics of symptomatic test-positive cases (by variants) and test-negative controls tested for SARS-CoV-2 between 14 December 2020 and 3 August 2021 in Ontario, Canada.

We identified 15,269 individuals with a COVID-19 hospitalization or death (Table 2). We observed largely the same sociodemographic patterns between individuals with severe outcomes caused by Delta versus those caused by other VOCs and non-VOC SARS-CoV-2.

**Table 2:** Characteristics of test-positive cases with severe outcomes (hospitalization or death) between 14 December 2020 and 3 August 2021 (based on test date) in Ontario, Canada.

| Characteristic | SARS-CoV-2-positive cases with severe outcomes (hospitalization or death) |  |  |  |  |  | SARS-CoV-2-negative symptomatic controls, n (%)<sup>*</sup> (N=598,905) |
| --- | --- | --- | --- | --- | --- | --- | --- |
|  | Alpha, n (%)<sup>*</sup> (N=7,438) | Beta, n (%)<sup>*</sup> (N=174) | Gamma, n (%)<sup>*</sup> (N=410) | Beta/Gamma, n (%)<sup>*</sup> (N=237) | Delta, n (%)<sup>*</sup> (N=639) | Non-VOC SARS-CoV-2, n (%)<sup>*</sup> (N=6,371) |  |
| BNT162b2 (Pfizer-BioNTech Comirnaty) |  |  |  |  |  |  |  |
| Received 1 dose only | 1,218 (16.4%) | 24 (13.8%) | 66 (16.1%) | 45 (19.0%) | 155 (24.3%) | 109 (1.7%) | 80,183 (13.4%) |
| Received 2 doses | 42 (0.6%) | ≤5 (≤2.9%) | ≤5 (≤1.2%) | ≤5 (≤2.1%) | 20 (3.1%) | ≤5 (≤0.1%) | 36,310 (6.1%) |
| mRNA-1273 (Moderna SpikeVax) |  |  |  |  |  |  |  |
| Received 1 dose only | 245 (3.3%) | ≤5 (≤2.9%) | 10 (2.4%) | 7 (3.0%) | 23 (3.6%) | 73 (1.1%) | 19,142 (3.2%) |
| Received 2 doses | 20 (0.3%) | 0 (0.0%) | 0 (0.0%) | ≤5 (≤2.1%) | ≤5 (≤0.8%) | ≤5 (≤0.1%) | 9,890 (1.7%) |
| ChAdOx1 (AstraZeneca Vaxzevria) |  |  |  |  |  |  |  |
| Received 1 dose only | 149 (2.0%) | ≤5 (≤2.9%) | 6 (1.5%) | 7 (3.0%) | 9 (1.4%) | ≤5 (≤0.1%) | 12,027 (2.0%) |
| Received 2 doses | ≤5 (≤0.1%) | 0 (0.0%) | 0 (0.0%) | 0 (0.0%) | ≤5 (≤0.8%) | 0 (0.0%) | 1,203 (0.2%) |
| Age (years), mean (standard deviation) | 60.5 (17.5) | 61.8 (15.6) | 62.8 (16.1) | 63.0 (15.4) | 57.2 (18.0) | 69.7 (17.3) | 43.2 (17.6) |
| Age group (years) |  |  |  |  |  |  |  |
| 16–29 | 377 (5.1%) | ≤5 (≤2.9%) | 11 (2.7%) | 9 (3.8%) | 45 (7.0%) | 171 (2.7%) | 160,115 (26.7%) |
| 30–39 | 618 (8.3%) | 11–15 (6.3–8.6%) | 24 (5.9%) | 8 (3.4%) | 71 (11.1%) | 283 (4.4%) | 125,797 (21.0%) |
| 40–49 | 938 (12.6%) | 22 (12.6%) | 45 (11.0%) | 25 (10.5%) | 96 (15.0%) | 403 (6.3%) | 101,635 (17.0%) |
| 50–59 | 1,529 (20.6%) | 33 (19.0%) | 94 (22.9%) | 50 (21.1%) | 148 (23.2%) | 767 (12.0%) | 95,615 (16.0%) |
| 60–69 | 1,529 (20.6%) | 46 (26.4%) | 88 (21.5%) | 55 (23.2%) | 104 (16.3%) | 1,150 (18.1%) | 64,675 (10.8%) |
| 70–79 | 1,317 (17.7%) | 34 (19.5%) | 80 (19.5%) | 53 (22.4%) | 94 (14.7%) | 1,434 (22.5%) | 32,223 (5.4%) |
| ≥80 | 1,130 (15.2%) | 23 (13.2%) | 68 (16.6%) | 37 (15.6%) | 81 (12.7%) | 2,163 (34.0%) | 18,845 (3.1%) |
| Male sex | 4,051 (54.5%) | 99 (56.9%) | 247 (60.2%) | 140 (59.1%) | 379 (59.3%) | 3,530 (55.4%) | 264,464 (44.2%) |
| Public health unit region<sup>†</sup> |  |  |  |  |  |  |  |
| Central East | 254 (3.4%) | ≤5 (≤2.9%) | 30 (7.3%) | 10 (4.2%) | 14 (2.2%) | 206 (3.2%) | 80,534 (13.4%) |
| Central West | 901 (12.1%) | 6 (3.4%) | 45 (11.0%) | 25 (10.5%) | 253 (39.6%) | 1,351 (21.2%) | 114,566 (19.1%) |
| Durham | 410 (5.5%) | ≤5 (≤2.9%) | 20 (4.9%) | ≤5 (≤2.1%) | 20 (3.1%) | 225 (3.5%) | 27,898 (4.7%) |
| Eastern | 158 (2.1%) | 12 (6.9%) | 6 (1.5%) | ≤5 (≤2.1%) | ≤5 (≤0.8%) | 120 (1.9%) | 22,461 (3.8%) |
| North | 163 (2.2%) | 7 (4.0%) | ≤5 (≤1.2%) | ≤5 (≤2.1%) | 13 (2.0%) | 203 (3.2%) | 49,571 (8.3%) |
| Ottawa | 397 (5.3%) | 65 (37.4%) | 8 (2.0%) | 8 (3.4%) | 11 (1.7%) | 208 (3.3%) | 5,719 (1.0%) |
| Peel | 957 (12.9%) | 8 (4.6%) | 70 (17.1%) | 19 (8.0%) | 82 (12.8%) | 552 (8.7%) | 63,469 (10.6%) |
| South West | 369 (5.0%) | ≤5 (≤2.9%) | 10 (2.4%) | 10 (4.2%) | 69 (10.8%) | 726 (11.4%) | 51,795 (8.6%) |
| Toronto | 3,168 (42.6%) | 58 (33.3%) | 179 (43.7%) | 136 (57.4%) | 156 (24.4%) | 2,181 (34.2%) | 119,674 (20.0%) |
| York | 625 (8.4%) | ≤5 (≤2.9%) | 35 (8.5%) | 20 (8.4%) | 16 (2.5%) | 577 (9.1%) | 60,235 (10.1%) |
| Biweekly period of test |  |  |  |  |  |  |  |
| 14 Dec 2020 to 27 Dec 2020 | 6 (0.1%) | 0 (0.0%) | 0 (0.0%) | 0 (0.0%) | 0 (0.0%) | 1,309 (20.5%) | 34,641 (5.8%) |
| 28 Dec 2020 to 10 Jan 2021 | 8 (0.1%) | 0 (0.0%) | 0 (0.0%) | 0 (0.0%) | 0 (0.0%) | 1,666 (26.1%) | 32,730 (5.5%) |
| 11 Jan 2021 to 24 Jan 2021 | 14 (0.2%) | 0 (0.0%) | ≤5 (≤1.2%) | 0 (0.0%) | 0 (0.0%) | 1,273 (20.0%) | 33,563 (5.6%) |
| 25 Jan 2021 to 7 Feb 2021 | 49 (0.7%) | ≤5 (≤2.9%) | ≤5 (≤1.2%) | 0 (0.0%) | 0 (0.0%) | 815 (12.8%) | 30,194 (5.0%) |
| 8 Feb 2021 to 21 Feb 2021 | 119 (1.6%) | ≤5 (≤2.9%) | 10 (2.4%) | 0 (0.0%) | 0 (0.0%) | 407 (6.4%) | 31,467 (5.3%) |
| 22 Feb 2021 to 7 Mar 2021 | 164 (2.2%) | 10 (5.7%) | 20 (4.9%) | ≤5 (≤2.1%) | 0 (0.0%) | 305 (4.8%) | 40,987 (6.8%) |
| 8 Mar 2021 to 21 Mar 2021 | 454 (6.1%) | 28 (16.1%) | 54 (13.2%) | 19 (8.0%) | 0 (0.0%) | 268 (4.2%) | 46,236 (7.7%) |
| 22 Mar 2021 to 4 Apr 2021 | 1,191 (16.0%) | 42 (24.1%) | 61 (14.9%) | 31 (13.1%) | 0 (0.0%) | 162 (2.5%) | 54,777 (9.1%) |
|  | Alpha, n (%)<sup>*</sup> (N=7,438) | Beta, n (%)<sup>*</sup> (N=174) | Gamma, n (%)<sup>*</sup> (N=410) | Beta/Gamma, n (%)<sup>*</sup> (N=237) | Delta, n (%)<sup>*</sup> (N=639) | Non-VOC SARS-CoV-2, n (%)<sup>*</sup> (N=6,371) |  |
| 5 Apr 2021 to 18 Apr 2021 | 2,052 (27.6%) | 50 (28.7%) | 117 (28.5%) | 75 (31.6%) | ≤5 (≤0.8%) | 119 (1.9%) | 67,854 (11.3%) |
| 19 Apr 2021 to 2 May 2021 | 1,618 (21.8%) | 29 (16.7%) | 91 (22.2%) | 41 (17.3%) | 15–19 (2.3–3.0%) | 18 (0.3%) | 62,310 (10.4%) |
| 3 May 2021 to 16 May 2021 | 1,016 (13.7%) | 7 (4.0%) | 35 (8.5%) | 25 (10.5%) | 80 (12.5%) | ≤5 (≤0.1%) | 46,541 (7.8%) |
| 17 May 2021 to 30 May 2021 | 457 (6.1%) | ≤5 (≤2.9%) | 14 (3.4%) | 32 (13.5%) | 98 (15.3%) | 0 (0.0%) | 29,745 (5.0%) |
| 31 May 2021 to 13 June 2021 | 203 (2.7%) | ≤5 (≤2.9%) | ≤5 (≤1.2%) | 7 (3.0%) | 114 (17.8%) | 16 (0.3%) | 22,497 (3.8%) |
| 14 June 2021 to 27 June 2021 | 64 (0.9%) | 0 (0.0%) | ≤5 (≤1.2%) | 0 (0.0%) | 118 (18.5%) | ≤5 (≤0.1%) | 20,625 (3.4%) |
| 28 June 2021 to 11 July 2021 | 18 (0.2%) | 0 (0.0%) | 0 (0.0%) | ≤5 (≤2.1%) | 94 (14.7%) | 7 (0.1%) | 15,819 (2.6%) |
| 12 July 2021 to 25 July 2021 | ≤5 (≤0.1%) | 0 (0.0%) | 0 (0.0%) | ≤5 (≤2.1%) | 69 (10.8%) | 0 (0.0%) | 17,649 (2.9%) |
| 26 July 2021 to 3 August 2021 | ≤5 (≤0.1%) | 0 (0.0%) | 0 (0.0%) | 0 (0.0%) | 46 (7.2%) | ≤5 (≤0.1%) | 11,270 (1.9%) |
| Number of tests in previous 3 months |  |  |  |  |  |  |  |
| 0 | 6,453 (86.8%) | 152 (87.4%) | 353 (86.1%) | 209 (88.2%) | 544 (85.1%) | 4,790 (75.2%) | 435,908 (72.8%) |
| 1 | 735 (9.9%) | 15 (8.6%) | 41 (10.0%) | 21 (8.9%) | 76 (11.9%) | 843 (13.2%) | 109,809 (18.3%) |
| ≥2 | 250 (3.4%) | 7 (4.0%) | 16 (3.9%) | 7 (3.0%) | 19 (3.0%) | 738 (11.6%) | 53,188 (8.9%) |
| Any comorbidity<sup>‡</sup> | 5,543 (74.5%) | 144 (82.8%) | 309 (75.4%) | 183 (77.2%) | 431 (67.4%) | 5,517 (86.6%) | 273,262 (45.6%) |
| Receipt of 2019-2020 and/or 2020-2021 influenza vaccination | 2,787 (37.5%) | 77 (44.3%) | 180 (43.9%) | 98 (41.4%) | 176 (27.5%) | 3,153 (49.5%) | 189,399 (31.6%) |
| Neighbourhood income quintile<sup>†, §</sup> |  |  |  |  |  |  |  |
| 1 (lowest) | 2,463 (33.1%) | 55 (31.6%) | 135 (32.9%) | 74 (31.2%) | 212 (33.2%) | 2,068 (32.5%) | 106,066 (17.7%) |
| 2 | 1,638 (22.0%) | 41 (23.6%) | 92 (22.4%) | 68 (28.7%) | 148 (23.2%) | 1,373 (21.6%) | 112,621 (18.8%) |
| 3 | 1,458 (19.6%) | 24 (13.8%) | 78 (19.0%) | 44 (18.6%) | 125 (19.6%) | 1,178 (18.5%) | 115,706 (19.3%) |
| 4 | 1,008 (13.6%) | 32 (18.4%) | 58 (14.1%) | 31 (13.1%) | 85 (13.3%) | 936 (14.7%) | 127,112 (21.2%) |
| 5 (highest) | 835 (11.2%) | 21 (12.1%) | 44 (10.7%) | 20 (8.4%) | 66 (10.3%) | 794 (12.5%) | 134,167 (22.4%) |
| Essential workers quintile<sup>†, ¶</sup> |  |  |  |  |  |  |  |
| 1 (0%–32.5%) | 1,171 (15.7%) | 50 (28.7%) | 54 (13.2%) | 37 (15.6%) | 79 (12.4%) | 992 (15.6%) | 107,842 (18.0%) |
| 2 (32.5%–42.3%) | 1,452 (19.5%) | 32 (18.4%) | 80 (19.5%) | 47 (19.8%) | 101 (15.8%) | 1,233 (19.4%) | 138,277 (23.1%) |
| 3 (42.3%–49.8%) | 1,452 (19.5%) | 30 (17.2%) | 84 (20.5%) | 50 (21.1%) | 131 (20.5%) | 1,249 (19.6%) | 124,149 (20.7%) |
| 4 (50.0%–57.5%) | 1,558 (20.9%) | 30 (17.2%) | 74 (18.0%) | 55 (23.2%) | 148 (23.2%) | 1,310 (20.6%) | 117,067 (19.5%) |
| 5 (57.5%–100%) | 1,767 (23.8%) | 31 (17.8%) | 115 (28.0%) | 48 (20.3%) | 176 (27.5%) | 1,554 (24.4%) | 106,993 (17.9%) |
| Persons per dwelling quintile<sup>†, **</sup> |  |  |  |  |  |  |  |
| 1 (0–2.1) | 1,327 (17.8%) | 44 (25.3%) | 60 (14.6%) | 52 (21.9%) | 128 (20.0%) | 1,470 (23.1%) | 96,399 (16.1%) |
| 2 (2.2–2.4) | 1,064 (14.3%) | 16 (9.2%) | 47 (11.5%) | 31 (13.1%) | 122 (19.1%) | 969 (15.2%) | 99,302 (16.6%) |
| 3 (2.5–2.6) | 795 (10.7%) | 26 (14.9%) | 54 (13.2%) | 34 (14.3%) | 72 (11.3%) | 733 (11.5%) | 76,071 (12.7%) |
| 4 (2.7–3.0) | 1,859 (25.0%) | 44 (25.3%) | 104 (25.4%) | 53 (22.4%) | 133 (20.8%) | 1,468 (23.0%) | 151,891 (25.4%) |
| 5 (3.1–5.7) | 2,351 (31.6%) | 43 (24.7%) | 142 (34.6%) | 67 (28.3%) | 180 (28.2%) | 1,691 (26.5%) | 170,522 (28.5%) |
| Self-identified visible minority quintile<sup>†, ††</sup> |  |  |  |  |  |  |  |
| 1 (0.0%–2.2%) | 401 (5.4%) | 10 (5.7%) | 25 (6.1%) | 18 (7.6%) | 74 (11.6%) | 527 (8.3%) | 97,243 (16.2%) |
| 2 (2.2%–7.5%) | 573 (7.7%) | 20 (11.5%) | 36 (8.8%) | 13 (5.5%) | 65 (10.2%) | 753 (11.8%) | 110,142 (18.4%) |
| 3 (7.5%–18.7%) | 922 (12.4%) | 26 (14.9%) | 49 (12.0%) | 27 (11.4%) | 93 (14.6%) | 1,090 (17.1%) | 109,977 (18.4%) |
| 4 (18.7%–43.5%) | 1,811 (24.3%) | 49 (28.2%) | 89 (21.7%) | 54 (22.8%) | 180 (28.2%) | 1,538 (24.1%) | 120,189 (20.1%) |
| 5 (43.5%–100%) | 3,693 (49.7%) | 68 (39.1%) | 208 (50.7%) | 125 (52.7%) | 223 (34.9%) | 2,430 (38.1%) | 156,787 (26.2%) |

<sup>\*</sup> Proportion reported, unless stated otherwise.
<sup>†</sup> The sum of counts does not equal the column total because of individuals with missing information (<1.0%) for this characteristic.
<sup>‡</sup>Comorbidities include chronic respiratory diseases, chronic heart diseases, hypertension, diabetes, immunocompromising conditions due to underlying diseases or therapy, autoimmune diseases, chronic kidney disease, advanced liver disease, dementia/frailty and history of stroke or transient ischemic attack.
<sup>§</sup>Neighbourhood income quintile has variable cut-off values in each city/Census area to account for cost of living. A dissemination area (DA) being in quintile 1 means it is among the lowest 20% of DAs in its city by income.
<sup>\*</sup>Percentage of people in the area working in the following occupations: sales and service occupations; trades, transport and equipment operators and related occupations; natural resources, agriculture, and related production occupations; and occupations in manufacturing and utilities. Census counts for people are randomly rounded up or down to the nearest number divisible by 5, which causes some minor imprecision.
<sup>\*\*</sup>Range of persons per dwelling.
<sup>††</sup>Percentage of people in the area who self-identified as a visible minority. Census counts for people are randomly rounded up or down to the nearest number divisible by 5, which causes some minor imprecision.

### Vaccine effectiveness against symptomatic infection

Against symptomatic infection caused by Alpha, vaccine effectiveness ≥14 days after the first dose was higher for mRNA-1273 (82%; 95% CI, 80–84%) than BNT162b2 (67%; 95% CI, 65– 68%) and ChAdOx1 (63%; 95% CI, 59–66%) (Figure 1a). Vaccine effectiveness was increased ≥7 days after the second dose against Alpha for all three vaccines: mRNA-1273=92% (95% CI, 88–95%), BNT162b2=89% (95% CI, 87–90%), and ChAdOx1=91% (95% CI, 62–98%).

**Figure 1:**
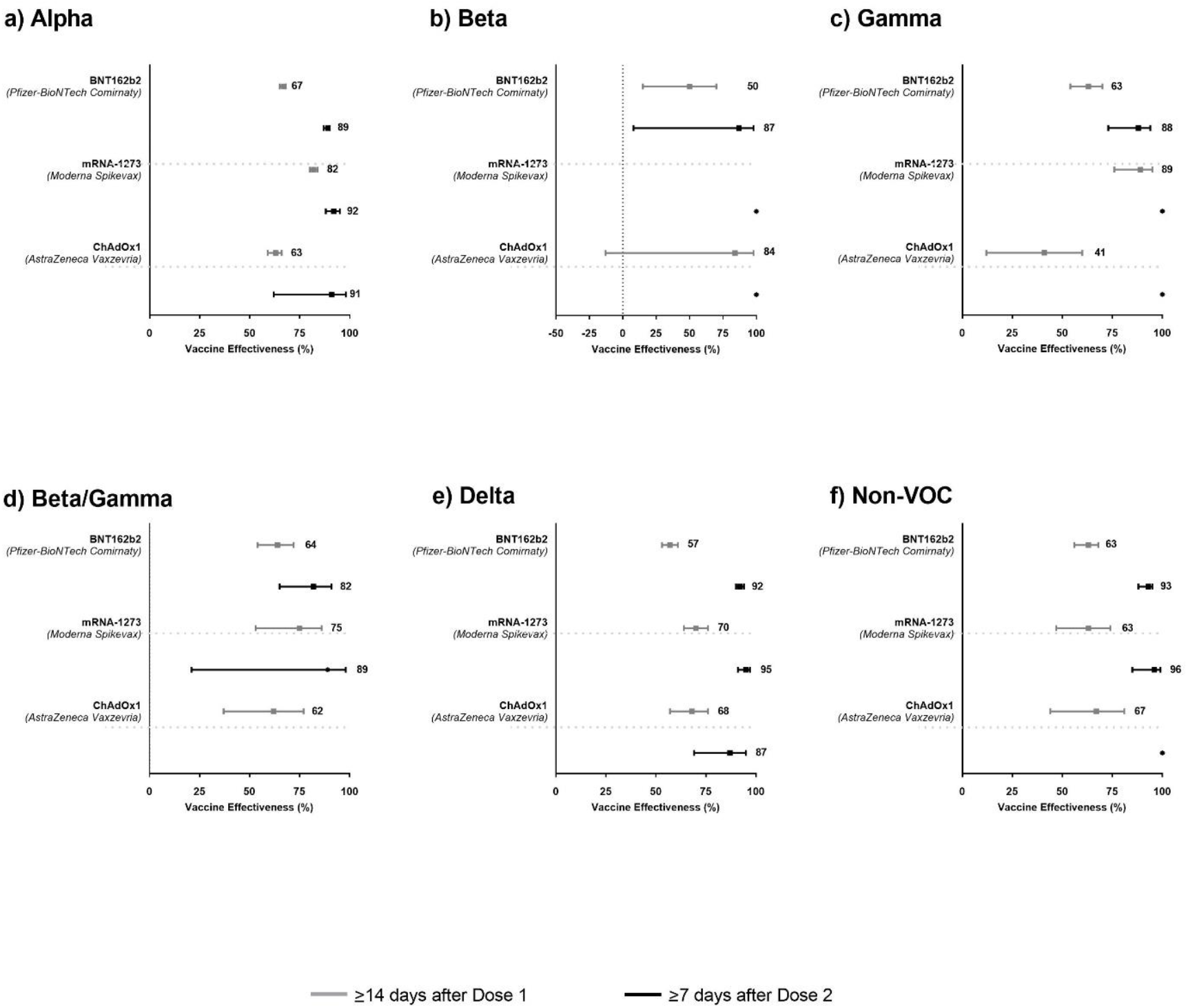
Adjusted vaccine effectiveness estimates of BNT162b2 (Pfizer-BioNTech Comirnaty), mRNA-1273 (Moderna Spikevax), and ChAdOx1 (AstraZeneca Vaxzevria) vaccines ≥14 days after the first dose (for individuals who received only one dose) and ≥7 days after the second dose against symptomatic SARS-CoV-2 infection caused by (a) Alpha, (b) Beta, (c) Gamma, (d) Beta/Gamma, (e) Delta, and (f) non-VOC SARS-CoV-2, between 14 December 2020 and 3 August 2021 in Ontario, Canada. Models were adjusted for age, sex, public health unit region, period of test (weekly period for Delta, and bi-weekly period for non-VOC SARS-CoV-2 and other VOCs), number of SARS-CoV-2 tests in the 3 months prior to 14 December 2020, presence of any comorbidity that increase the risk of severe COVID-19, receipt of 2019/2020 and/or 2020/2021 influenza vaccination, and Census dissemination area-level quintiles of household income, proportion of persons employed as non-health essential workers, persons per dwelling, and proportion of self-identified visible minorities. Data are presented as effectiveness point estimates, with error bars indicating the corresponding 95% CIs. Solid circles indicate vaccine effectiveness estimated as 100% based on zero vaccinated test-positive cases.

Estimates of effectiveness against symptomatic infection caused by Beta (Figure 1b) were imprecise or were 100% because of low numbers, or absence, of vaccinated test-positive cases, respectively (Supplementary Table 1). Vaccine effectiveness against symptomatic infection caused by Gamma was higher after the first dose for mRNA-1273 (89%; 95% CI, 76– 95%) than ChAdOx1 (41%; 95% CI, 12–60%), and was intermediate for BNT162b2 (63%; 95% CI, 54–70%) (Figure 1c). Effectiveness against Gamma increased after the second dose for BNT162b2 but could not be reliably estimated for mRNA-1273 and ChAdOx1 because of zero vaccinated cases. Protection against symptomatic infection caused by Beta/Gamma appeared to be marginally higher (but with overlapping confidence intervals) after the first dose for mRNA-1273 (75%; 95% CI, 53–86%) than BNT162b2 (64%; 95% CI, 54–72%) and ChAdOx1 (62%; 95% CI, 37–77%) (Figure 1d). Receipt of the second dose increased vaccine effectiveness against Beta/Gamma for BNT162b2 (82%; 95% CI, 65–91%) and mRNA-1273 (89%; 95% CI, 21–98%) but could not be reliably estimated for ChAdOx1 due to an absence of any vaccinated cases (Supplementary Table 1).

Against Delta, vaccine effectiveness after the first dose was higher for mRNA-1273 (70%; 95% CI, 64–76%) and ChAdOx1 (68%; 95% CI, 57–76%) than BNT162b2 (57%; 95% CI, 53–61%) (Figure 1e). Vaccine effectiveness was increased after the second dose for all three vaccines, to 95% (95% CI, 91–97%) for mRNA-1273, 87% (95% CI, 69–95%) for ChAdOx1, and 92% (95% CI, 90–94%) for BNT162b2.

By product, vaccine effectiveness after one dose tended to be lower against Delta than against Alpha and Gamma for mRNA-1273 (70% vs. 82% and 89%) and for BNT162b2 (57% vs. 67% and 63%), but was similar to Alpha for ChAdOx1 (68% vs. 63%). Two doses of BNT162b2 and mRNA-1273 increased protection against Delta (92–95%) to levels comparable to Alpha (89–92%), Beta (87%), and Gamma (88%).

### Vaccine effectiveness against hospitalization or death

Vaccine effectiveness against hospitalization or death caused by all four VOCs was generally higher than against symptomatic infection after the first dose for all three vaccines (Figure 2). In particular, against Delta, vaccine effectiveness against severe outcomes after the first dose of BNT162b2, mRNA-1273, and ChAdOx1 was 81% (95% CI, 76–85%), 90% (95% CI, 82–94%), and 91% (95% CI, 72–97%), respectively (Figure 2e). Receipt of the second dose was associated with vaccine effectiveness estimates above 90% against: Alpha, Beta, and Delta for BNT162b2; Alpha and Delta for mRNA-1273; and Delta for ChAdOx1. Estimates were in the 80% range against Gamma for BNT162b2 and Alpha for ChAdOx1, and could not be reliably estimated for other VOC-vaccine combinations due to low numbers, or absence, of vaccinated cases.

**Figure 2:**
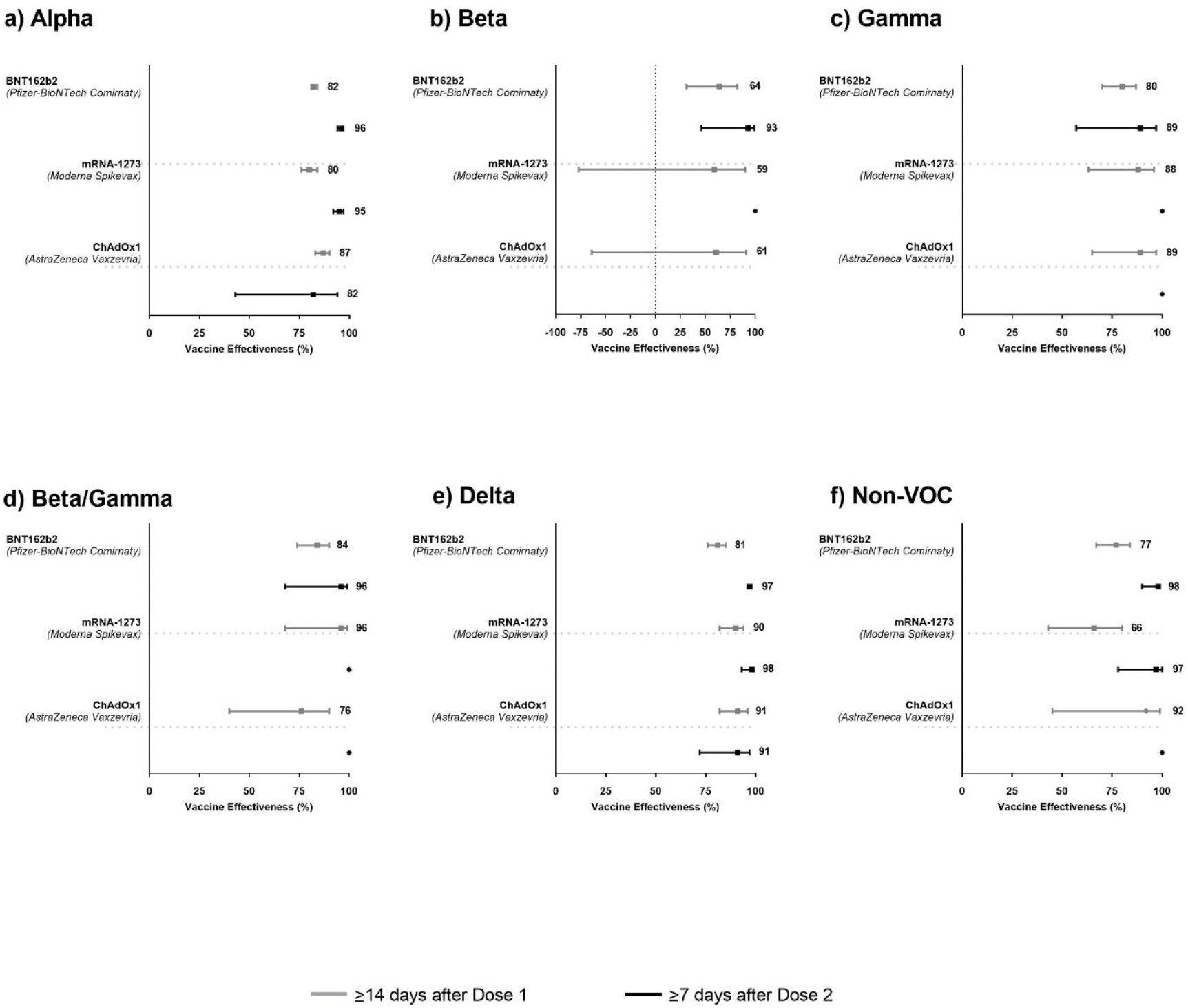
Adjusted vaccine effectiveness estimates of BNT162b2 (Pfizer-BioNTech Comirnaty), mRNA-1273 (Moderna Spikevax), and ChAdOx1 (AstraZeneca Vaxzevria) vaccines ≥14 days after the first dose (for individuals who received only one dose) and ≥7 days after the second dose against severe outcomes (hospitalization or death) caused by (a) Alpha, (b) Beta, (c) Gamma, (d) Beta/Gamma, (e) Delta, and (f) non-VOC SARS-CoV-2, between 14 December 2020 and 17 August 2021 in Ontario, Canada. Models were adjusted for age, sex, public health unit region, period of test (weekly period for Delta, and bi-weekly period for non-VOC SARS-CoV-2 and other VOCs), number of SARS-CoV-2 tests in the 3 months prior to 14 December 2020, presence of any comorbidity that increase the risk of severe COVID-19, receipt of 2019/2020 and/or 2020/2021 influenza vaccination, and Census dissemination area-level quintiles of household income, proportion of persons employed as non-health essential workers, persons per dwelling, and proportion of self-identified visible minorities. Data are presented as effectiveness point estimates, with error bars indicating the corresponding 95% CIs. Solid circles indicate vaccine effectiveness estimated as 100% based on zero vaccinated test-positive cases.

### Sensitivity analyses

In sensitivity analyses, vaccine effectiveness estimates using ≥21 days after the first dose and ≥14 days after the second dose were mostly very similar (estimates differed by ≤5%) to our primary analyses (Table 3). Where differences were >5%, the sensitivity analyses yielded higher estimates. Similar vaccine effectiveness estimates were also observed when limiting the study period to 05 April 2021 to 3 August 2021 (Supplementary Table 2).

**Table 3:** Vaccine effectiveness against Alpha (B.1.1.7), Beta (B.1.351), Gamma (P.1), Beta/Gamma, and Delta (B.1.617.2) variants of concern by outcome, vaccine product, number of doses received, and time between most recent vaccination date and index date for those tested for SARS-CoV-2 between 14 December 2020 and 3 August 2021 in Ontario, Canada.

| Outcome | Vaccine effectiveness* (95% CI) |  |  |  |  |  |
| --- | --- | --- | --- | --- | --- | --- |
|  | Alpha | Beta | Gamma | Beta/Gamma† | Delta‡ | Non-VOC SARS-CoV-2 |
| **Symptomatic infection** |  |  |  |  |  |  |
| **BNT162b2 (Pfizer-BioNTech Comirnaty)§** |  |  |  |  |  |  |
| ≥14 days after one dose only | 67 (65, 68) | 50 (15, 70) | 63 (54, 70) | 64 (54, 72) | 57 (53, 61) | 63 (56, 68) |
| ≥21 days after one dose only | 70 (69, 72) | 48 (6, 71) | 67 (57, 74) | 68 (57, 76) | 59 (54, 63) | 65 (58, 71) |
| ≥7 days after two doses | 89 (87, 90) | 87 (8, 98) | 88 (73, 94) | 82 (65, 91) | 92 (90, 94) | 93 (88, 95) |
| ≥14 days after two doses | 88 (86, 90) | 86 (0, 98) | 90 (76, 96) | 89 (74, 96) | 92 (89, 94) | 92 (87, 95) |
| **mRNA-1273 (Moderna SpikeVax)§** |  |  |  |  |  |  |
| ≥14 days after one dose only | 82 (80, 84) | -** | 89 (76, 95) | 75 (53, 86) | 70 (64, 76) | 63 (47, 74) |
| ≥21 days after one dose only | 83 (80, 85) | -** | 90 (73, 96) | 74 (48, 87) | 69 (62, 75) | 70 (51, 81) |
| ≥7 days after two doses | 92 (88, 95) | -¶ | -¶ | 89 (21, 98) | 95 (91, 97) | 96 (85, 99) |
| ≥14 days after two doses | 92 (87, 95) | -¶ | -¶ | 88 (10, 98) | 94 (90, 97) | 98 (83, 100) |
| **ChAdOx1 (AstraZeneca Vaxzevria)§** |  |  |  |  |  |  |
| ≥14 days after one dose only | 63 (59, 66) | 84 (-13, 98) | 41 (12, 60) | 62 (37, 77) | 68 (57, 76) | 67 (44, 81) |
| ≥21 days after one dose only | 71 (67, 74) | 79 (-53, 97) | 46 (13, 66) | 59 (29, 76) | 68 (57, 76) | 80 (56, 91) |
| ≥7 days after two doses | 91 (62, 98) | -¶ | -¶ | -** | 87 (69, 95) | -¶ |
| ≥14 days after two doses | 87 (47, 97) | -¶ | -¶ | -** | 88 (68, 96) | -¶ |
| **Hospitalization or death** |  |  |  |  |  |  |
| **BNT162b2 (Pfizer-BioNTech Comirnaty)§** |  |  |  |  |  |  |
| ≥14 days after one dose only | 82 (81, 84) | 64 (31, 82) | 80 (70, 87) | 84 (74, 90) | 81 (76, 85) | 77 (67, 84) |
| ≥21 days after one dose only | 87 (85, 88) | 65 (23, 84) | 88 (79, 93) | 87 (77, 92) | 81 (76, 85) | 88 (79, 94) |
| ≥7 days after two doses | 96 (94, 97) | 93 (46, 99) | 89 (57, 97) | 96 (68, 99) | 97 (96, 98) | 98 (90, 99) |
| ≥14 days after two doses | 96 (94, 97) | 92 (39, 99) | 94 (59, 99) | 95 (64, 99) | 98 (96, 99) | 97 (88, 99) |
| **mRNA-1273 (Moderna SpikeVax)§** |  |  |  |  |  |  |
| ≥14 days after one dose only | 80 (76, 84) | 59 (-77, 90) | 88 (63, 96) | 96 (68, 99) | 90 (82, 94) | 66 (43, 80) |
| ≥21 days after one dose only | 82 (77, 86) | -** | 95 (63, 99) | -¶ | 91 (83, 95) | 70 (41, 85) |
| ≥7 days after two doses | 95 (92, 97) | -¶ | -¶ | -¶ | 98 (93, 99) | 97 (78, 100) |
| ≥14 days after two doses | 95 (92, 97) | -¶ | -¶ | -¶ | 98 (93, 100) | -¶ |
| **ChAdOx1 (AstraZeneca Vaxzevria)§** |  |  |  |  |  |  |
| ≥14 days after one dose only | 87 (83, 90) | 61 (-64, 91) | 89 (65, 97) | 76 (40, 90) | 91 (82, 96) | 92 (45, 99) |
| ≥21 days after one dose only | 91 (88, 94) | 73 (-98, 96) | 86 (57, 96) | 72 (28, 89) | 91 (81, 95) | 90 (27, 99) |
| ≥7 days after two doses | 82 (43, 94) | -** | -¶ | -¶ | 91 (72, 97) | -¶ |
| ≥14 days after two doses | 92 (41, 99) | -** | -¶ | -¶ | 90 (67, 97) | -¶ |

\*Adjusted for age, sex, public health unit region, period of test (weekly period for Delta, and bi-weekly period for non-VOC SARS-CoV-2 and other VOCs), number of SARS-CoV-2 tests in the 3 months prior to 14 December 2020, presence of any comorbidity that increase the risk of severe COVID-19, receipt of 2019/2020 and/or 2020/2021 influenza vaccination, and Census dissemination area-level quintiles of household income, proportion of persons employed as non-health essential workers, persons per dwelling, and proportion of self-identified visible minorities
†RT-PCR testing dates for both test-positive cases and test-negative controls were restricted to 11 January 2021 to 3 August 2021 for Beta, Gamma, and Beta/Gamma.
<sup>‡</sup>RT-PCR testing dates for both test-positive cases and test-negative controls were restricted to 5 April 2021 to 3 August 2021 for Delta.
<sup>§</sup>For one dose only, excludes individuals who received two doses; shorter intervals after the doses include the longer interval (i.e., analyses for $\geq 14$ days after the first dose also includes subjects who were $\geq 21$ days after the first dose, and analyses for $\geq 7$ days also includes subjects who were $\geq 14$ days after the second dose).
<sup>¶</sup>VE estimated as 100% based on zero vaccinated test-positive cases.
<sup>\*\*</sup>VE not reported due to extremely imprecise 95% confidence intervals.

### Vaccine effectiveness by age group

In age group-stratified analyses, vaccine effectiveness against symptomatic infection caused by all VOCs was in most instances lower or similar in older adults (aged ≥60 years) compared to younger individuals (aged <60 years) after partial vaccination (Table 4). Vaccine effectiveness in older adults increased to levels comparable to younger individuals after the second dose, except against Gamma for BNT162b2 and against Delta for ChAdOx1. Vaccine effectiveness was higher against hospitalization and death than symptomatic infection for both older and younger adults.

**Table 4.** Vaccine effectiveness against Alpha (B.1.1.7), Beta (B.1.351), Gamma (P.1), Beta/Gamma, and Delta (B.1.617.2) variants of concern by outcome, vaccine product, number of doses received, and age group for those tested for SARS-CoV-2 between 14 December 2020 and 3 August 2021 in Ontario, Canada.

| Outcome | Vaccine effectiveness* (95% CI) |  |  |  |  |  |
| --- | --- | --- | --- | --- | --- | --- |
|  | Alpha | Beta<sup>†</sup> | Gamma<sup>†</sup> | Beta/Gamma<sup>†</sup> | Delta<sup>‡</sup> | Non-VOC SARS-CoV-2 |
| **Symptomatic infection** |  |  |  |  |  |  |
| **BNT162b2 (Pfizer-BioNTech Comirnaty)** |  |  |  |  |  |  |
| ≥14 days after one dose only<sup>§</sup> |  |  |  |  |  |  |
| <60 years | 72 (70, 73) | 50 (4, 74) | 72 (61, 79) | 62 (49, 72) | 60 (55, 64) | 65 (58, 71) |
| ≥60 years | 60 (56, 63) | 43 (-49, 78) | 45 (21, 62) | 68 (49, 80) | 37 (18, 52) | 60 (46, 70) |
| ≥7 days after two doses |  |  |  |  |  |  |
| <60 years | 88 (85, 90) | 83 (-22, 98) | 93 (78, 98) | 82 (60, 92) | 92 (90, 94) | 95 (90, 97) |
| ≥60 years | 91 (87, 94) | -<sup>¶</sup> | 58 (-23, 86) | 79 (4, 95) | 89 (83, 93) | 85 (69, 93) |
| **mRNA-1273 (Moderna SpikeVax)** |  |  |  |  |  |  |
| ≥14 days after one dose only<sup>§</sup> |  |  |  |  |  |  |
| <60 years | 84 (81, 86) | -<sup>**</sup> | 88 (70, 95) | 81 (56, 91) | 74 (68, 79) | 73 (55, 83) |
| ≥60 years | 79 (74, 83) | -<sup>**</sup> | 94 (53, 99) | 65 (10, 86) | 48 (20, 67) | 48 (12, 69) |
| ≥7 days after two doses |  |  |  |  |  |  |
| <60 years | 89 (83, 93) | -<sup>¶</sup> | -<sup>¶</sup> | 84 (-19, 98) | 96 (92, 98) | 97 (77, 100) |
| ≥60 years | 95 (89, 98) | -<sup>**</sup> | -<sup>¶</sup> | -<sup>¶</sup> | 92 (77, 97) | 95 (67, 99) |
| **ChAdOx1 (AstraZeneca Vaxzevria)** |  |  |  |  |  |  |
| ≥14 days after one dose only<sup>§</sup> |  |  |  |  |  |  |
| <60 years | 65 (60, 70) | -<sup>**</sup> | 36 (-12, 64) | 75 (39, 90) | 68 (56, 77) | 77 (29, 93) |
| ≥60 years | 63 (57, 68) | -<sup>¶</sup> | 48 (7, 71) | 53 (8, 76) | 64 (35, 80) | 69 (43, 83) |
| ≥7 days after two doses |  |  |  |  |  |  |
| <60 years | -<sup>¶</sup> | -<sup>¶</sup> | -<sup>¶</sup> | -<sup>**</sup> | 94 (55, 99) | -<sup>¶</sup> |
| ≥60 years | 88 (49, 97) | -<sup>**</sup> | -<sup>¶</sup> | -<sup>¶</sup> | 81 (44, 93) | -<sup>¶</sup> |
| **Hospitalization or death** |  |  |  |  |  |  |
| **BNT162b2 (Pfizer-BioNTech Comirnaty)** |  |  |  |  |  |  |
| ≥14 days after one dose only<sup>§</sup> |  |  |  |  |  |  |
| <60 years | 92 (90, 94) | -<sup>¶</sup> | 90 (60, 98) | -<sup>¶</sup> | 89 (83, 92) | 84 (62, 93) |
| ≥60 years | 81 (79, 83) | 52 (0, 77) | 80 (69, 87) | 80 (66, 88) | 74 (65, 81) | 75 (62, 83) |
| ≥7 days after two doses |  |  |  |  |  |  |
| <60 years | 99 (94, 100) | -<sup>**</sup> | -<sup>¶</sup> | -<sup>¶</sup> | 99 (96, 100) | -<sup>¶</sup> |
| ≥60 years | 95 (92, 96) | 89 (13, 99) | 87 (45, 97) | 95 (62, 99) | 96 (93, 98) | 96 (83, 99) |
| **mRNA-1273 (Moderna SpikeVax)** |  |  |  |  |  |  |
| ≥14 days after one dose only<sup>§</sup> |  |  |  |  |  |  |
| <60 years | 91 (85, 95) | -<sup>**</sup> | -<sup>¶</sup> | 77 (-74, 97) | 94 (84, 98) | 88 (10, 98) |
| ≥60 years | 77 (71, 81) | 77 (-77, 97) | 87 (57, 96) | -<sup>¶</sup> | 85 (71, 92) | 59 (30, 76) |
| ≥7 days after two doses |  |  |  |  |  |  |
| <60 years | -<sup>¶</sup> | -<sup>**</sup> | -<sup>¶</sup> | -<sup>¶</sup> | -<sup>¶</sup> | -<sup>¶</sup> |
|  | Alpha | Beta<sup>†</sup> | Gamma<sup>†</sup> | Beta/Gamma<sup>†</sup> | Delta<sup>‡</sup> | Non-VOC SARS-CoV-2 |
| ≥60 years | 94 (91, 97) | -<sup>¶</sup> | -<sup>¶</sup> | -<sup>¶</sup> | 96 (88, 99) | 96 (71, 99) |
| **ChAdOx1 (AstraZeneca Vaxzevria)** |  |  |  |  |  |  |
| ≥14 days after one dose only<sup>§</sup> |  |  |  |  |  |  |
| <60 years | 85 (77, 90) | -<sup>¶</sup> | -<sup>¶</sup> | -<sup>¶</sup> | 94 (81, 98) | 74 (-95, 97) |
| ≥60 years | 88 (85, 91) | -<sup>**</sup> | 88 (61, 96) | 65 (10, 87) | 88 (69, 95) | -<sup>¶</sup> |
| ≥7 days after two doses |  |  |  |  |  |  |
| <60 years | -<sup>**</sup> | -<sup>**</sup> | -<sup>**</sup> | -<sup>**</sup> | 75 (-4, 94) | -<sup>¶</sup> |
| ≥60 years | 84 (34, 96) | -<sup>**</sup> | -<sup>¶</sup> | -<sup>¶</sup> | 96 (70, 99) | -<sup>¶</sup> |

\*Adjusted for age, sex, public health unit region, period of test (weekly period for Delta, and bi-weekly period for non-VOC SARS-CoV-2 and other VOCs), number of SARS-CoV-2 tests in the 3 months prior to 14 December 2020, presence of any comorbidity that increase the risk of severe COVID-19, receipt of 2019/2020 and/or 2020/2021 influenza vaccination, and Census dissemination area-level quintiles of household income, proportion of persons employed as non-health essential workers, persons per dwelling, and proportion of self-identified visible minorities.
<sup>†</sup>RT-PCR testing dates for both test-positive cases and test-negative controls were restricted to 11 January 2021 to 3 August 2021 for Beta, Gamma, and Beta/Gamma.
<sup>‡</sup>RT-PCR testing dates for both test-positive cases and test-negative controls were restricted to 5 April 2021 to 3 August 2021 for Delta.
<sup>§</sup>Excludes individuals who received two doses.
<sup>¶</sup>VE estimated as 100% based on zero vaccinated test-positive cases.
<sup>\*\*</sup>VE not reported due to extremely imprecise 95% confidence intervals.

## Discussion

We estimated that one dose of BNT162b2 and mRNA-1273 was >50% and >70% effective, respectively, against symptomatic infection caused by VOCs that have circulated to date in Ontario, Canada. One dose of ChAdOx1 prevented 41% of symptomatic infections by Gamma, and was >60% effective against Alpha and Delta. For all three vaccines, effectiveness increased (to >80% and, in most cases, >90%) following the second dose. Effectiveness of the first dose was substantially higher against hospitalization or death than against symptomatic infection for all VOC-vaccine combinations except for mRNA-1273 against Alpha and Gamma because vaccine effectiveness against symptomatic infection was already high; the second dose of mRNA vaccines further improved effectiveness against severe outcomes. We also found that assessing effectiveness ≥21 days after the first dose and ≥14 days after the second dose resulted in similar or higher estimates, restricting the analysis to a period when all VOCs were co-circulating yielded similar results, and the effectiveness of one dose tended to be lower for older adults compared to younger adults.

Our vaccine effectiveness estimates against symptomatic COVID-19 infection with Alpha and Gamma after the first dose of mRNA or ChAdOx1 vaccines are similar to estimates against infection from British Columbia, Canada (67% for mRNA vaccines against Alpha and 61% against Gamma)^15^ and Qatar (88% for mRNA-1273 against Alpha)^9^ but higher than estimates after one dose against infection or symptomatic COVID-19 reported from Qatar (for BNT162b2, 30% against Alpha and 17% against Beta^7^; 61% for mRNA-1273 against Beta^9^), England (49% for BNT162b2 and 51% for ChAdOx1 against Alpha)^10^ and Scotland (27% for BNT162b2 and 39% for ChAdOx1 against Alpha)^6^. Our estimates after one dose are also higher than the estimates for BNT162b2 against severe, critical, or fatal disease in Qatar (54%)^7^, but comparable with the estimates for hospitalization in England (83%)^8^ caused by the Alpha variant with BNT162b2 or ChAdOx1 and for severe, critical, or fatal disease with mRNA-1273 caused by Alpha/Beta in Qatar (82%)^9^.

We estimated higher vaccine effectiveness against symptomatic COVID-19 infection with the Delta variant after one dose of mRNA or ChAdOx1 vaccines than the effectiveness against symptomatic or asymptomatic infections reported from England and Scotland (30–33% for BNT162b2 and 18–33% for ChAdOx1)^6 10^. However, our effectiveness estimates against symptomatic infections with Delta after the first dose were lower than the estimates against infections in Qatar (64% for BNT162b2 and 79% for mRNA-1273)^11^. Our estimates after the second dose were higher than the estimates against symptomatic infections reported in England (88% for BNT162b2 and 67% for ChAdOx1)^10^, Scotland (83% for BNT162b2 and 61% for ChAdOx1)^6^, Israel (40.5% for BNT162b2)^16^, the USA (42% for BNT162b2 and 76% for mRNA-1273 against infection)^17^ and Qatar (53.5% for BNT162b2 and 84.8% for mRNA-1273 against infection)^11^. Against hospitalization with the Delta variant, our vaccine effectiveness after one dose was lower for BNT162b2 and higher for ChAdOx1 than the estimates from England (81% vs. 94% for BNT162b2 and 91% vs. 71% for ChAdOx1)^8^. Vaccine effectiveness against severe, critical and fatal COVID-19 disease with Delta after one dose of BNT162b2 and mRNA-1273 in Qatar was reported to be 100% because of zero vaccinated cases^11^.

After the second dose, our estimates against both outcomes for all VOCs were higher than after one dose, and comparable with estimates reported in previous studies^6-8 10^. However, the vaccine effectiveness after two doses was reported to be lower than after one dose against any infection with Delta (54% vs. 64%) in Qatar, which the authors interpreted as waning of protection^11^.

The heterogeneity in vaccine estimates, particularly after one dose, across studies could result from a number of factors, including differences in study design, study population, SARS-CoV-2 test assays and testing criteria, comprehensiveness of test results recorded in databases, outcome definitions and ascertainment, timing of VOC circulation, vaccine priority groups, vaccine rollout, interval between vaccine doses, and variables adjusted to control for possible confounding^18^.

Province-wide data allowed us to estimate the effectiveness of all three vaccines used in Canada against symptomatic infection and severe outcomes caused by the four VOCs that have circulated in Canada thus far. The test-negative study design has the advantage to control for bias resulting from differences in healthcare-seeking behaviour between vaccinated and unvaccinated individuals^19^.

There are some limitations of our study. First, VOC classification in this study relied on a combination of mutation screening and whole genome sequencing, and the criteria for sequencing evolved over time. Our definition of Delta specimens initially relied largely on a proxy measure of a N501Y-/E484K-result on mutation screening and a combination of date and geographic location, which were used to infer probable Delta variant specimens. Thus, a small proportion of specimens classified as Delta may have been non-VOC SARS-CoV-2 specimens. Second, since vaccine effectiveness is likely impacted by age, interval between vaccine doses, time between vaccine receipt and index date, vaccine product, and VOC, and given that the eligibility criteria for vaccination (e.g., initial prioritization of older age groups), availability of certain vaccine products, and distribution of circulating VOCs all varied over time, comparisons of vaccine effectiveness estimates between combinations of vaccine products and VOCs should be made with caution. However, we included a sensitivity analysis that restricted the study period to individuals tested during 5 April through 3 August 2021 (to ensure that all VOCs and non-VOC SARS-CoV-2 were circulating and to mitigate temporal confounding caused by the aforementioned factors), which yielded very similar results to our primary analysis. Third, it is possible that we may have under-ascertained severe outcomes if they were not recorded in the public health surveillance database used, such as when severe outcomes occurred after completion of case follow-up or when case volumes exceeded public health system capacity and public health investigation of each laboratory-confirmed case was not possible. This may have resulted in overestimation of vaccine effectiveness against severe outcomes. Fourth, we used specimen collection date as the index date because of lack of available data on symptom onset date in OLIS, which precluded us from restricting the study population to individuals who were tested within 10 days of symptom onset. Thus, we may have underestimated vaccine effectiveness by increasing the risk of false-negative cases by extending the interval between symptom onset and testing. Last, despite our best efforts to adjust for potential confounders and the use of the test-negative design, these results may nonetheless be susceptible to residual confounding given the observational nature of the study.

In conclusion, our real-world vaccine effectiveness estimates suggest that even a single dose of these three COVID-19 vaccine products provide considerable protection against symptomatic infection and severe outcomes caused by these four VOCs, particularly for young adults, and that two doses provide even higher protection. While vaccine effectiveness estimates after two doses are presently most relevant to high-income countries with adequate vaccine supply and relatively higher two-dose vaccine coverage, effectiveness estimates after one dose remain relevant for low- and middle-income countries with suboptimal vaccine supply and low one- or two-dose vaccine coverage. Thus, our findings have public health policy implications worldwide. Jurisdictions facing COVID-19 vaccine supply constraints may benefit from delaying the second dose (at least for younger individuals) to maximize the number of individuals receiving partial protection from the first dose, thereby potentially providing greater overall protection of the population more rapidly. However, older adults would likely benefit most from minimizing the delay in receiving the second dose to achieve adequate protection against VOCs, including Delta, the predominant VOC currently in circulation worldwide.

## Methods

We employed a test-negative design to compare vaccination status between test-positive individuals (with symptomatic infection or a severe outcome) and symptomatic but test-negative individuals^19^. We included community-dwelling Ontarians aged ≥16 years who had symptoms consistent with or a severe outcome attributable to COVID-19, and who were tested for SARS-CoV-2 between 14 December 2020 and 3 August 2021. We excluded individuals who had tested positive for SARS-CoV-2 prior to their selected index date and those who received Ad26.COV2.S (Janssen), because it has not been used in Ontario (despite receiving approval), or mixed (ChAdOx1-mRNA or different mRNA) vaccine schedules.

### Data sources and definitions

Comprehensive province-wide datasets for SARS-CoV-2 laboratory testing, SARS-CoV-2 public health surveillance, COVID-19 vaccination, and healthcare system use were linked using unique encoded identifiers and analyzed at ICES (formerly the Institute for Clinical Evaluative Sciences). Details have been described previously^20^.

#### Vaccination status

We obtained information regarding COVID-19 vaccination status, including vaccine product, date of administration, and dose number, from COVaxON, a centralized COVID-19 vaccine information system in Ontario.

#### COVID-19 testing and identification of variants

Data on laboratory-confirmed SARS-CoV-2 infection detected by real-time reverse transcription polymerase chain reaction (RT-PCR) were collected from the Ontario Laboratories Information System (OLIS) for both individuals who tested positive (treated as cases) and individuals who tested negative (treated as controls). We used specimen collection date as the index date because symptom onset date was not consistently available in OLIS. We used the first positive test for cases, and a randomly selected negative test for controls with multiple negative tests during the study period.

We obtained information on variants from the Public Health Case and Contact Management system (CCM), which contains results of screening tests for mutations and whole genome sequencing to assign SARS-CoV-2 lineage or variant of concern (VOC). All RT-PCR positive specimens with cycle threshold values ≤35 were screened for N501Y and E484K mutations by multiplex RT-PCR (VOC PCR)^21^. From 7 June 2021, specimens positive for both N501Y and E484K mutations were also screened for K417N and K417T mutations to differentiate specimens between Beta and Gamma variants^21^.

At the beginning of 2021, whole genome sequencing was performed on specimens that had specific mutations detected by VOC PCR to confirm they were indeed VOCs. From 3 February 2021, specimens with the N501Y mutation, and from 22 March 2021, all specimens with the E484K mutation and 5% of E484K-negative specimens (all with cycle threshold values ≤30) were sequenced for surveillance purposes^21 22^. A subset of RT-PCR-positive specimens without any mutations detected by VOC PCR were also selected for sequencing for surveillance purposes^21^. Additionally, VOC PCR testing and sequencing were performed for specific indications such as recent travellers, partially or fully vaccinated individuals, cases of suspected reinfection, or to support investigations of outbreaks and potential super-spreading events^23^. Ontario started sequencing 10% and 50% of VOC PCR-screened specimens on 2 May 2021 and 30 May 2021, respectively. From 14 June 2021, Ontario started sequencing 100% of eligible VOC PCR-screened specimens^21^ that continued until the end of the study period before scaling back to sequencing 50% of specimens on 27 August 2021^22^.

In addition to those classified into SARS-CoV-2 lineages based on sequencing, we considered specimens positive for the N501Y mutation and negative for the E484K mutation (N501Y+/E484K-) as Alpha. Both Beta and Gamma have N501Y and E484K mutations, and can only be differentiated by additional screening for K417N and K417T mutations. Hence, we considered specimens positive for N501Y, E484K, and K417N mutations as Beta, and specimens positive for N501Y, E484K, and K417T as Gamma. We grouped the specimens that could not be separated into Beta and Gamma into a combined Beta/Gamma group. We classified specimens collected after 1 April 2021 that were negative for both N501Y and E484K (N501Y-/E484K-) mutations as either probable, possible, or unlikely Delta cases based on the predicted probability that it was Delta. To do this, we created a logistic regression model of the probability a N501Y-/E484K-case was Delta based on the date of specimen collection and the forward sortation area (geographical unit based on the first three characters of the postal code) ranked by the cumulative incidence of laboratory-confirmed SARS-CoV-2 cases between 23 January 2020 and 28 March 2021 and grouped into deciles^24^. For each decile, we examined the trajectories of the daily counts of N501Y-/E484K-specimens between 1 April 2021 and 30 May 2021 to estimate the predicted probability that a N501Y-/E484K-specimen represented Delta. We classified specimens with >75% probability of being a Delta case to be ‘probable Delta’ cases, those with 25-75% probability to be ‘possible Delta’ cases, and those with <25% probability to be ‘unlikely Delta’ cases. Our approach correlates well with sequencing results (n=538) for the province indicating a rapid increase in the proportion of N501Y-/E484K-cases being identified as Delta from mid-March to mid-May 2021^25^. As of 31 May 2021, all N501Y-/E484K-specimens were considered Delta^22^. We grouped the probable Delta cases with those identified through sequencing. We classified specimens with no lineage information and N501Y-/E484K-specimens collected prior to 1 April 2021 as non-VOC SARS-CoV-2. We also grouped the ‘unlikely Delta’ cases with the non-VOC specimens. We excluded ‘possible Delta’ cases and N501Y-/E484K+ cases from our analyses.

#### Outcomes

For vaccine effectiveness against symptomatic infection, individuals who were symptomatic and tested positive for SARS-CoV-2 in OLIS were considered as cases. For severe outcomes, test-positive individuals who had a hospitalization or death up to 17 August 2021 (regardless of the presence of any symptoms recorded at the time of RT-PCR testing) were identified from CCM and considered as cases. Individuals who were symptomatic but only had tests negative for SARS-CoV-2 in OLIS were considered as controls for both outcomes. However, for severe outcomes, we excluded symptomatic test-negative individuals who later tested positive between 4 August 2021 and 17 August 2021.

#### Covariates

We obtained information on the following covariates from administrative databases: age and sex from the Ontario Registered Persons Database (RPDB); postal code and Public Health Unit of residence from the RPDB and Statistics Canada Postal Code Conversion File Plus (version 7B); the number of SARS-CoV-2 RT-PCR tests for each individual during the 3 months prior to 14 December (a proxy for individuals who are at increased risk of exposure to SARS-CoV-2 infection and undergo frequent testing), and biweekly (weekly for Delta) period of RT-PCR test to account for the temporal viral activity and regional vaccine roll-out created using testing information from OLIS; comorbidities^26^ associated with increased risk of severe COVID-19 identified from various databases using validated algorithms and commonly used diagnostic codes and algorithms described previously^27^, including Expanded Diagnostic Clusters and Special Population Markers from the Johns Hopkins ACG System (version 10)^28^; influenza vaccination status during the 2019/2020 and/or 2020/2021 influenza season (a proxy for health behaviours) determined from physician and pharmacist billing claims in the Ontario Health Insurance Plan and Ontario Drug Benefit databases, respectively; and neighbourhood-level information on median household income, proportion of the working population employed as non-health essential workers, average number of persons per dwelling, and proportion of the population who self-identify as a visible minority obtained from 2016 Census data. Details regarding these covariates are provided in Supplementary Table 3^20^.

### Statistical analyses

We used multivariable logistic regression models to estimate the odds ratio comparing the odds of vaccination in test-positive cases with the odds of vaccination among test-negative controls, adjusting for the aforementioned covariates that are associated with COVID-19 and vaccine uptake^26 29 30^. We calculated vaccine effectiveness using the following formula: Vaccine effectiveness = 1 – (odds ratio) x 100%.

We estimated vaccine effectiveness against SARS-CoV-2 infection and severe COVID-19 outcomes (hospitalization or death) caused by non-VOC SARS-CoV-2, Alpha, Beta, Gamma, Beta/Gamma, and Delta separately by vaccine product (BNT162b2, mRNA-1273, and ChAdOx1) and number of doses received. For individuals who had received only one dose by the index date, we calculated vaccine effectiveness ≥14 days after the first dose. For individuals who had received two doses, we calculated vaccine effectiveness ≥7 days after the second dose. As a sensitivity analysis, and to facilitate comparisons with other studies, we also estimated vaccine effectiveness ≥21 days after the first dose and ≥14 days after the second dose.

When estimating vaccine effectiveness against Beta, Gamma, Beta/Gamma and Delta, we restricted both test-positive cases and test-negative controls to those who were tested on/after the dates of initial confirmation of these variants in Ontario (11 January 2021 for Beta, Gamma, and Beta/Gamma; 5 April 2021 for Delta). Furthermore, since the primary periods of circulation for non-VOC SARS-CoV-2 and VOCs varied relative to the vaccination campaign (i.e., more non-VOC SARS-CoV-2 circulated earlier in the campaign when fewer individuals were vaccinated, whereas Delta circulated later when more were vaccinated), we conducted a sensitivity analysis restricted to individuals who were tested between 5 April 2021 and 3 August 2021 when non-VOC SARS-CoV-2 and VOCs were concurrently circulating, thereby accounting for differences in vaccine availability and coverage over time.

Lastly, we estimated vaccine effectiveness stratified by age group (<60 years and ≥60 years).

All analyses were conducted using SAS Version 9.4 (SAS Institute Inc., Cary, NC). All tests were two-sided and used p*<*0.05 as the level of statistical significance. We did not report estimates of vaccine effectiveness when 95% confidence intervals [CIs] were extremely imprecise (i.e., ranging between a very large negative number and nearly 100) or when vaccine effectiveness was estimated as 100% based on zero vaccinated test-positive cases and the 95% CIs were essentially infinite.

## Data Availability

The dataset from this study is held securely in coded form at ICES. While legal data sharing agreements between ICES and data providers (e.g., healthcare organizations and government) prohibit ICES from making the dataset publicly available, access may be granted to those who meet pre-specified criteria for confidential access, available at www.ices.on.ca/DAS. The full dataset creation plan and underlying analytic code are available from the authors upon request, understanding that the computer programs may rely upon coding templates or macros that are unique to ICES and are therefore either inaccessible or may require modification.

## Correspondence and requests for materials

should be addressed to J.C.K.

## Acknowledgments

We would like to acknowledge Public Health Ontario for access to case-level data from CCM and COVID-19 laboratory data, as well as assistance with data interpretation. We also thank the staff of Ontario’s public health units who are responsible for COVID-19 case and contact management and data collection within CCM. We thank IQVIA Solutions Canada Inc. for use of their Drug Information Database. The authors are grateful to the Ontario residents without whom this research would be impossible.

## Author contributions

J.C.K. and H.C. designed and oversaw the study. S.N., S.H. and H.C. obtained the data and conducted all analyses (data set and variable creation and statistical modelling). B.C. contributed to data analyses and data preparation for the symptomatic data set. H.C. and S.H. verified the data in the study. S.N. did the literature search. S.N. and J.C.K. drafted the manuscript. All authors contributed to the analysis plan, interpreted the results, critically reviewed and edited the manuscript, approved the final version, and agreed to be accountable for all aspects of the work.

## Ethics approval

ICES is a prescribed entity under Ontario’s Personal Health Information Protection Act (PHIPA). Section 45 of PHIPA authorizes ICES to collect personal health information, without consent, for the purpose of analysis or compiling statistical information with respect to the management of, evaluation or monitoring of, the allocation of resources to or planning for all or part of the health system. Projects that use data collected by ICES under section 45 of PHIPA, and use no other data, are exempt from REB review. The use of the data in this project is authorized under section 45 and approved by ICES’ Privacy and Legal Office.

## Disclaimers

This study was supported by ICES, which is funded by an annual grant from the Ontario Ministry of Health (MOH) and the Ministry of Long-Term Care (MLTC). This study was supported by the Ontario Health Data Platform (OHDP), a Province of Ontario initiative to support Ontario’s ongoing response to COVID-19 and its related impacts. The study sponsors did not participate in the design and conduct of the study; collection, management, analysis and interpretation of the data; preparation, review or approval of the manuscript; or the decision to submit the manuscript for publication. Parts of this material are based on data and/or information compiled and provided by the Canadian Institute for Health Information (CIHI) and by Cancer Care Ontario (CCO). However, the analyses, conclusions, opinions and statements expressed herein are solely those of the authors, and do not reflect those of the funding or data sources; no endorsement by ICES, MOH, MLTC, OHDP, its partners, the Province of Ontario, CIHI or CCO is intended or should be inferred.

