## Supplementary material for "Effectiveness of mRNA and ChAdOx1 COVID-19 vaccines against symptomatic SARS-CoV-2 infection and severe outcomes with variants of concern in Ontario"

**Supplementary Table 1.** Counts of test-positive cases and test-negative controls among those tested for SARS-CoV-2 between 14 December 2020 and 3 August 2021 in Ontario, Canada

| **Outcome** | **Alpha** | | **Beta*** | | **Gamma*** | |
|---|---|---|---|---|---|---|
| | **Test-positive cases, No. Vaccinated/ Total‡** | **Test-negative controls, No. Vaccinated/ Total‡** | **Test-positive cases, No. Vaccinated/ Total‡** | **Test-negative controls, No. Vaccinated/ Total‡** | **Test-positive cases, No. Vaccinated/ Total‡** | **Test-negative controls, No. Vaccinated/ Total‡** |
| **Symptomatic infection** | | | | | | |
| **BNT162b2 (Pfizer-BioNTech Comirnaty)** | | | | | | |
| ≥14 days after one dose only§ | 2,484 / 40,105 | 60,775 / 501,166 | 17 / 348 | 60,758 / 434,122 | 104 / 1,782 | 60,758 / 434,122 |
| ≥7 days after two doses | 138 / 37,759 | 32,292 / 472,683 | -¶ | 32,292 / 405,656 | -¶ | 32,292 / 405,656 |
| **mRNA-1273 (Moderna SpikeVax)** | | | | | | |
| ≥14 days after one dose only§ | 282 / 37,903 | 13,750 / 454,141 | -¶ | 13,750 / 387,114 | 6 / 1,684 | 13,750 / 387,114 |
| ≥7 days after two doses | -¶ | 8,608 / 448,999 | 0 / 331 | 8,608 / 381,972 | 0 / 1,678 | 8,608 / 381,972 |
| **ChAdOx1 (AstraZeneca Vaxzevria)** | | | | | | |
| ≥14 days after one dose only§ | 496 / 38,117 | 8,976 / 449,367 | -¶ | 8,976 / 382,340 | 27 / 1,705 | 8,976 / 382,340 |
| ≥7 days after two doses | -¶ | 1,096 / 441,487 | 0 / 331 | 1,096 / 374,460 | 0 / 1,678 | 1,096 / 374,460 |
| **Hospitalization or death** | | | | | | |
| **BNT162b2 (Pfizer-BioNTech Comirnaty)** | | | | | | |
| ≥14 days after one dose only§ | 637 / 6,398 | 60,766 / 500,916 | 12 / 155 | 60,749 / 433,899 | 33 / 359 | 60,749 / 433,899 |
| ≥7 days after two doses | 28 / 5,789 | 32,277 / 472,427 | -¶ | 32,277 / 405,427 | -¶ | 32,277 / 405,427 |
| **mRNA-1273 (Moderna SpikeVax)** | | | | | | |
| ≥14 days after one dose only§ | 117 / 5,878 | 13,746 / 453,896 | -¶ | 13,746 / 386,896 | -¶ | 13,746 / 386,896 |
| ≥7 days after two doses | -¶ | 8,605 / 448,755 | 0 / 143 | 8,605 / 381,755 | 0 / 326 | 8,605 / 381,755 |
| **ChAdOx1 (AstraZeneca Vaxzevria)** | | | | | | |
| ≥14 days after one dose only§ | 73 / 5,834 | 8,973 / 449,123 | -¶ | 8,973 / 382,123 | -¶ | 8,973 / 382,123 |
| ≥7 days after two doses | -¶ | 1,095 / 441,245 | 0 / 143 | 1,095 / 374,245 | 0 / 326 | 1,095 / 374,245 |
| | **Beta/Gamma*** | | **Delta†** | | **Non-VOC SARS-CoV-2** | |
| | **Test-positive cases, No. Vaccinated/ Total‡** | **Test-negative controls, No. Vaccinated/ Total‡** | **Test-positive cases, No. Vaccinated/ Total‡** | **Test-negative controls, No. Vaccinated/ Total‡** | **Test-positive cases, No. Vaccinated/ Total‡** | **Test-negative controls, No. Vaccinated/ Total‡** |

| | | | | | | |
|---|---|---|---|---|---|---|
| **Symptomatic infection** | | | | | | |
| **BNT162b2 (Pfizer-BioNTech Comirnaty)** | | | | | | |
| ≥14 days after one dose only[§] | 78 / 1,126 | 60758 / 434,122 | 657 / 2,489 | 54770 / 210,002 | 189 / 30,861 | 60,775 / 501,166 |
| ≥7 days after two doses | -[¶] | 32292 / 405,656 | 87 / 1,919 | 28335 / 183,567 | 18 / 30,690 | 32,292 / 472,683 |
| **mRNA-1273 (Moderna SpikeVax)** | | | | | | |
| ≥14 days after one dose only[§] | 11 / 1,059 | 13750 / 387,114 | 117 / 1,949 | 12955 / 168,187 | 32 / 30,704 | 13,750 / 454,141 |
| ≥7 days after two doses | -[¶] | 8608 / 381,972 | 14 / 1,846 | 7807 / 163,039 | -[¶] | 8,608 / 448,999 |
| **ChAdOx1 (AstraZeneca Vaxzevria)** | | | | | | |
| ≥14 days after one dose only[§] | 16 / 1,064 | 8976 / 382,340 | 57 / 1,889 | 8737 / 163,969 | 14 / 30,686 | 8,976 / 449,367 |
| ≥7 days after two doses | -[¶] | 1096 / 374,460 | -[¶] | 1096 / 156,328 | -[¶] | 1,096 / 441,487 |
| **Hospitalization or death** | | | | | | |
| **BNT162b2 (Pfizer-BioNTech Comirnaty)** | | | | | | |
| ≥14 days after one dose only[§] | 23 / 198 | 60749 / 433,899 | 122 / 546 | 54761 / 209,872 | 34 / 6,214 | 60,766 / 500,916 |
| ≥7 days after two doses | -[¶] | 32277 / 405,427 | -[¶] | 28321 / 183,432 | -[¶] | 32,277 / 472,427 |
| **mRNA-1273 (Moderna SpikeVax)** | | | | | | |
| ≥14 days after one dose only[§] | -[¶] | 13746 / 386,896 | 14 / 438 | 12951 / 168,062 | 16 / 6,196 | 13,746 / 453,896 |
| ≥7 days after two doses | 0 / 175 | 8605 / 381,755 | -[¶] | 7804 / 162,915 | -[¶] | 8,605 / 448,755 |
| **ChAdOx1 (AstraZeneca Vaxzevria)** | | | | | | |
| ≥14 days after one dose only[§] | -[¶] | 8973 / 382,123 | -[¶] | 8734 / 163,845 | -[¶] | 8,973 / 449,123 |
| ≥7 days after two doses | 0 / 175 | 1095 / 374,245 | -[¶] | 1095 / 156,206 | 0 / 6,180 | 1,095 / 441,245 |

[*]RT-PCR testing dates for both test-positive cases and test-negative controls were restricted to 11 January 2021 to 3 August 2021 for Beta, Gamma, and Beta/Gamma.

[†]RT-PCR testing dates for both test-positive cases and test-negative controls were restricted to 5 April 2021 to 3 August 2021 for Delta.

[‡]The numerator represents the number of test-positive cases or test-negative controls that are vaccinated, and the denominator represents the total number of vaccinated and unvaccinated test-positive cases or test-negative controls.

[§]Excludes individuals who received two doses.

[¶]Counts suppressed because they are either small cell counts (≤5, which cannot be disclosed because of privacy and data obligations) or they could be derived using other numbers in this table or other tables.

**Supplementary Table 2.** Vaccine effectiveness against Alpha (B.1.1.7), Beta (B.1.351), Gamma (P.1), and Beta/Gamma variants of concern by outcome, vaccine product, and number of doses received for those tested for SARS-CoV-2 between 5 April 2021 and 3 August 2021 in Ontario, Canada

| **Outcome** | **Vaccine effectiveness* (95% CI)** | | | | |
|---|---|---|---|---|---|
| | **Alpha** | **Beta** | **Gamma** | **Beta/Gamma** | **Non-VOC SARS-CoV-2** |
| **Symptomatic infection** | | | | | |
| **BNT162b2 (Pfizer-BioNTech Comirnaty)** | | | | | |
| ≥14 days after one dose only[§] | 69 (67, 70) | 46 (3, 70) | 66 (57, 73) | 65 (54, 73) | 63 (54, 70) |
| ≥7 days after two doses | 89 (87, 91) | 81 (-38, 97) | 89 (74, 95) | 82 (63, 91) | 91 (82, 95) |
| **mRNA-1273 (Moderna SpikeVax)** | | | | | |
| ≥14 days after one dose only[§] | 83 (81, 85) | -[**] | 90 (77, 95) | 75 (55, 87) | 62 (41, 75) |
| ≥7 days after two doses | 92 (88, 95) | -[**] | -[¶] | 89 (21, 99) | 93 (70, 98) |
| **ChAdOx1 (AstraZeneca Vaxzevria)** | | | | | |
| ≥14 days after one dose only[§] | 66 (62, 69) | 81 (-42, 97) | 43 (14, 62) | 64 (39, 78) | 81 (59, 91) |
| ≥7 days after two doses | 91 (65, 98) | -[¶] | -[¶] | -[**] | -[¶] |
| **Hospitalization or death** | | | | | |
| **BNT162b2 (Pfizer-BioNTech Comirnaty)** | | | | | |
| ≥14 days after one dose only[§] | 86 (84, 87) | 72 (37, 87) | 86 (78, 91) | 86 (76, 91) | 83 (70, 91) |
| ≥7 days after two doses | 96 (94, 97) | 88 (6, 99) | 90 (60, 98) | 95 (65, 99) | 95 (79, 99) |
| **mRNA-1273 (Moderna SpikeVax)** | | | | | |
| ≥14 days after one dose only[§] | 84 (80, 87) | -[**] | 93 (73, 98) | 96 (69, 99) | 83 (44, 95) |
| ≥7 days after two doses | 96 (93, 98) | -[¶] | -[¶] | -[¶] | -[¶] |
| **ChAdOx1 (AstraZeneca Vaxzevria)** | | | | | |
| ≥14 days after one dose only[§] | 88 (85, 91) | 61 (-70, 91) | 93 (73, 98) | 77 (40, 91) | 91 (35, 99) |
| ≥7 days after two doses | 83 (47, 95) | -[**] | -[**] | -[¶] | -[¶] |

[*]Adjusted for age, sex, public health unit region, week of test, number of SARS-CoV-2 tests in the 3 months prior to 14 December 2020, presence of any comorbidity that increase the risk of severe COVID-19, receipt of 2019/2020 and/or 2020/2021 influenza vaccination, and Census dissemination area-level quintiles of household income, proportion of persons employed as non-health essential workers, persons per dwelling, and proportion of self-identified visible minorities.
[§]Excludes individuals who received two doses.
[¶]VE estimated as 100% based on zero vaccinated test-positive cases.
[**]VE not reported due to extremely imprecise 95% confidence intervals.

**Supplementary Table 3.** Description of covariates

| Variable | Definition |
| --- | --- |
| Age | Age was determined from the Registered Persons Database. This variable was included *a priori* as hypothesized to be directly related to COVID-19 infection risk.[1] |
| Sex | Sex was determined from the Registered Persons Database. This variable was included *a priori* as hypothesized to be directly related to COVID-19 infection risk.[1] |
| Biweekly period of COVID-19 test | 14 Dec 2020 to 27 Dec 2020<br>28 Dec 2020 to 10 Jan 2021<br>11 Jan 2021 to 24 Jan 2021<br>25 Jan 2021 to 7 Feb 2021<br>8 Feb 2021 to 21 Feb 2021<br>22 Feb 2021 to 7 Mar 2021<br>8 Mar 2021 to 21 Mar 2021<br>22 Mar 2021 to 4 Apr 2021<br>5 Apr 2021 to 18 Apr 2021<br>19 Apr 2021 to 2 May 2021<br>3 May 2021 to 16 May 2021<br>17 May 2021 to 30 May 2021<br>31 May 2021 to 13 June 2021<br>14 June 2021 to 27 June 2021<br>28 June 2021 to 11 July 2021<br>12 July 2021 to 25 July 2021<br>26 July 2021 to 3 August 2021 |
| Geographic unit (Public health unit regions in Ontario) | Taken from Public Health Unit (PHU) information using postal code of residence as recorded in the Registered Persons Database and Statistics Canada Postal Code Conversion File Plus (version 7B). Regions were defined as follows:<br>Central East: PHU 35 (Haliburton, Kawartha, Pine Ridge District Health Unit), 55 (Peterborough County—City Health Unit), 60 (Simcoe Muskoka District Health Unit)<br>Central West: PHU 27 (Brant County Health Unit), 34 (Haldimand-Norfolk Health Unit), 36 (Halton Regional Health Unit), 37 (City of Hamilton Health Unit), 46 (Niagara Regional Area Health Unit), 65 (Waterloo Health Unit), 66 (Wellington-Dufferin-Guelph Health Unit)<br>Durham: PHU 30 (Durham Regional Health Unit)<br>Eastern: PHU 38 (Hastings and Prince Edward Counties Health Unit), 41 (Kingston, Frontenac and Lennox and Addington Health Unit), 43 (Leeds, Grenville and Lanark District Health Unit), 57 (Renfrew County and District Health Unit), 58 (The Eastern Ontario Health Unit)<br>North: PHU 26 (The District of Algoma Health Unit), 47 (North Bay Parry Sound District Health Unit), 49 (Northwestern Health Unit), 56 (Porcupine Health Unit), 61 (Sudbury and District Health Unit), 62 (Thunder Bay District Health Unit), 63 (Timiskaming Health Unit)<br>Ottawa: PHU 51 (City of Ottawa Health Unit)<br>Peel: PHU 53 (Peel Regional Health Unit)<br>South West: PHU 31 (Elgin-St. Thomas), 33 (Grey Bruce Health Unit), 39 (Huron County Health Unit), 40 (Chatham-Kent Health Unit), 42 (Lambton Health Unit), 44 (Middlesex-London Health Unit), 52 (Oxford), 54 (Perth District Health Unit), 68 (Windsor-Essex County Health Unit), 75 (Southwestern Health Unit)<br>Toronto: PHU 95 (City of Toronto Health Unit)<br>York: PHU 70 (York Regional Health Unit) |
| Chronic heart disease | Individuals were defined as having "chronic heart disease" if they had congestive heart failure (CHF), ischemic heart disease, or atrial fibrillation. The definitions for these conditions are as follows:<br><br>CHF:[2]<br>An ICES-derived CHF database was used to identify patients with CHF, based on 1 NACRS, DAD, SDS, or OHIP claim and a second claim (from either) in 1 year. The CHF database is limited to those aged 40 years or older. This variable was included a priori as hypothesized to be directly related to COVID-19 infection risk.[3]<br><br>OHIP: 428<br>DAD, SDS: ICD-9: 428, ICD-10: I500, I501, I509<br><br>Cardiac ischemic disease:[4]<br>Any comorbidity in the past 5 years (DAD, any diagnosis field) or history of procedure in past 20 years (DAD, SDS), of the following:<br><br>Comorbidity (DAD, any diagnosis in the past 5 years): |

| Variable | Definition |
|---|---|
| | Angina: ICD-10: I20<br><br>Chronic Ischemic Heart Disease: ICD-10: I25;<br>Myocardial infarction: ICD-10: I21, I22<br><br>Procedure (DAD & SDS):<br>Coronary Artery Bypass Grafting:<br>CCI procedure codes: 1IJ76<br>CCP procedure codes: 481<br><br>Percutaneous Coronary Intervention:<br>CCI procedure codes: 1IJ50, 1IJ54, 1IJ57GQ<br>CCP procedure codes: 4802, 4803<br><br>Atrial fibrillation:[5]<br>Individuals with 1 hospitalization or 4 MD visits within a year in the past 5 years with the following codes:<br>ICD-9: 427.31, 427.32<br>ICD-10: I48<br>OHIP dxcode: 427 |
| Hypertension | An ICES-specific hypertension database[6] was used to identify patients with hypertension, based on 1 or more DAD diagnoses or 2 or more OHIP diagnoses in a two-year period; or 1 OHIP diagnosis followed by an OHIP/DAD diagnosis within two years. This variable was included *a priori* as hypothesized to be directly related to COVID-19 infection risk.[3]<br><br>DAD, SDS: ICD-9: 401, 402, 403, 404, 405; ICD-10: I10, I11, I12, I13, I15<br><br>OHIP diagnostic codes: 401, 402, 403, 404, or 405 |
| Diabetes | An ICES-specific diabetes database[7] was used to identify patients with diabetes, based on 2 OHIP diagnostic codes or 1 OHIP service code or 1 DAD admission within 2 years. This variable was included *a priori* as hypothesized to be directly related to COVID-19 infection risk.[3]<br><br>DAD, SDS: ICD-9: 250; ICD-10: E10, E11, E13, E14<br>OHIP: 250<br>OHIP service codes: Q040, K029, K030, K045, K046 |
| Asthma | An ICES-specific asthma database was used to identify patients with asthma, based on 2 or more ambulatory care visits and/or 1 or more hospitalizations. This variable was included a priori as hypothesized to be directly related to COVID-19 infection risk, as a result of its relationship to severe COVID-19 outcomes.[8]<br><br>OHIP<br>OHIP diagnostic code: 493<br><br>DAD<br>ICD-9 diagnostic code: 493<br>ICD-10 diagnostic codes: J45, J46 |
| Chronic obstructive pulmonary disease (COPD) | An ICES-specific COPD database was used to identify patients with COPD, based on 1 or more ambulatory care visits and/or 1 or more hospitalizations. The algorithm to identify COPD patients was only validated in those aged 35 years or older. This variable was included a priori as hypothesized to be directly related to COVID-19 infection risk.[3]<br><br>OHIP<br>OHIP diagnostic codes: 491, 492, 496<br><br>DAD<br>ICD-9 diagnostic codes: 491, 492, 496<br>ICD-10 diagnostic codes: J41, J42, J43, J44 |
| Immunocompromised (HIV, transplant, immunosuppressive therapy) | We included immunosuppressive conditions *a priori* as hypothesized to be directly related to COVID-19 infection risk.[3]<br><br>HIV:[9]<br>An ICES-specific HIV database was used to identify patients with HIV, based on 3 physician claims in 3 years with OHIP diagnostic codes: 042, 043 or 044 |

| Variable | Definition |
|---|---|
| | Solid organ transplant recipients:<br>CORRLINK is an ICES-specific database which links CORR (Canadian Organ Replacement Register) and DAD data. This database only includes patients that have received an organ transplant and does not include dialysis patients.<br>• For transplants before December 31, 2019: individuals are a transplant recipient if they have a treatment code of 171, where the treatment was before the index date<br>• For transplants on/after January 1, 2020: Identify ICD-10 codes, CCI procedure codes, and OHIP feecodes from DAD, NACRS, and OHIP (codes available upon request)<br><br>Allogenic/autologous bone marrow transplant recipients:<br>We identified those who had a history of allogenic bone marrow transplant before the index date using the following combination of diagnostic codes:<br>DAD:<br>• CCP procedure codes = 53.0<br>• CCI procedure codes = 1WY19, 1LZ19HHU7, 1LZ19HHU8<br>OHIP:<br>• Feecode = Z426<br><br>Other immune disorders:<br>Individuals were identified as having disorders of the immune system based on health care encounters recorded in DAD, SDS, NACRS, and OHIP in the 2-years prior to index using the Johns Hopkins ACG ® System Version 10.[10]<br><br>Any hospitalization (any diagnosis field) with the following codes:<br>• Sickle-cell disease (ICD-10 D57.0 – D57.2; D57.8 OR ICD-9 282.6);<br>• Other immune system disorders (ICD-9 273.2, 279.0, 279.1, 279.2, 279.3, 279.8, 279.9, 289.8; ICD-10 D80, D81, D82, D83, D84, D89; OHIP dxcode 279)<br>• Immunosuppressive therapy (>30 days (total days supplied) of oral corticosteroid in the 6 months before index date; receipt of other immunocompromising drug, including antineoplastics, in the 6 months before index date)<br><br>Active cancer:<br>• Any of the following treatments in the past 6 months: cancer surgery (codes available upon request), radiation (if the ICD-10 code listed was Z510 in NACRS), chemotherapy (if the ICD-10 code listed was Z511 or Z512 and any evidence of cancer diagnosis in the Ontario Cancer Registry (OCR) prior to the last treatment date)<br>• If not any of the above, individuals still classified as having cancer if they had a cancer diagnosis in OCR in the year before the index date |
| Autoimmune disease | Individuals considered to have autoimmune disease if they had any of the following:<br>• Rheumatoid arthritis[11,12] (identified in the Ontario Rheumatoid Arthritis Database)<br>• Inflammatory bowel disease (identified in the Ontario Crohn's and Colitis Cohort)[13,14]<br>• Psoriasis/psoriatic arthritis[15]<br> o Psoriasis: 1 hospitalization or 3 physician billings prior to index date. DAD: ICD-9: 696.1, 696.8; ICD-10: L40.0, L40.1, L40.2, L40.3, L40.4, L40.8, L40.9. OHIP: dxcode = 696<br> o Psoriatic arthritis: 1 hospitalization or: (3 physician billings for psoriatic arthritis + 1 billing for psoriasis [696]). DAD: ICD-9: 696.0; ICD-10: L40.5, M07.0, M07.1, M07.2, M07.3, M09.0. OHIP: dxcode = 721 (at least one of these billings must be billed by a rheumatologist).<br>• Multiple sclerosis[16]<br> o Individuals with one hospitalization or 5 physician billings over 2 years. DAD: ICD-9: 340; ICD-10: G35. OHIP: dxcode = 340. |
| Chronic kidney disease (CKD) | This variable was included *a priori* as hypothesized to be directly related to COVID-19 infection risk.[3] We defined this variable as having a CKD diagnosis code in DAD, NACRS, OHIP in the past 5 years, or: at least 1 dialysis code in each of the 3 months prior to index [17]<br><br>OHIP: 403, 585<br><br>ICD-10: E102, E112, E132, E142, I12, I13, N08, N18, N19 |
| | Patients who were on chronic dialysis in the year before index date, identified as those with at least 2 of any of the following codes in OHIP, DAD, or SDS separated by at least 90 days, but less than 150 days:[18] |

| Variable | Definition |
|---|---|
| | OHIP service codes: R849, G323, G325, G326, G860, G862, G865 G863, G866, G330, G331, G332, G333, G861, G082, G083, G085, G090, G091, G092, G093, G094, G095, G096, G294, G295, G864, H540, H740<br><br>DAD, SDS:<br>CCI procedure codes: 5195, 6698<br>CCP procedure code: 1PZ21 |
| Advanced liver disease (Cirrhosis or Decompensated Cirrhosis) | We included advanced liver disease *a priori* as hypothesized to be directly related to COVID-19 infection risk.[3]<br><br>Defined using the Cirrhosis Algorithm 9 [from [19]]:<br>Two or more physician visits (diagnosis code 571), or<br>one or more hospital diagnosis of cirrhosis, using the following diagnostic codes:<br>ICD-9 : 456.1, 571.2, 571.5<br>ICD-10: I85.9, I98.2, K70.3,K71.7, K74.6<br><br>Defined using the Decompensated Cirrhosis Algorithm 5 (from above reference):<br>One or more physician visits with diagnosis code 571 and (one or more hospital diagnosis or one or more procedure), using the following diagnostic codes:<br><br>ICD-9: 456.0, 456.2, 572.2, 572.3, 572.4, 782.4, 789.5l; ICD-10: I85.0, I86.4, I98.20, I98.3, K721, K729, K76.6, K76.7, R17, R18<br><br>CCI: 1.NA.13.BA-FA, 1.NA.13.BA-X7, 1.NA.13.BA-BD, 1.KQ.76GP-NR, 1.OT.52.HA<br><br>CCP: 1006, 6691<br><br>OHIP: J057, Z591 |
| Dementia | Dementia (ICES cohort) definition:<br>1 hospitalization for dementia and/or 3 ambulatory visits for dementia, each separated by at least 30 days, within 2 years and/or 1 prescription from ODB.[20] This variable was included *a priori* as hypothesized to be directly related to COVID-19 infection risk,[3] as well as a marker for healthcare access, mobility, and household-level exposures.[21 22]<br><br>OHIP: 290, 331<br><br>DAD, SDS:<br>ICD-9: 0461, 290.0, 290.1, 290.2, 290.3, 290.4, 294, 331.0, 331.1, 331.5<br>ICD-10: F00, F01, F02, F03, G30<br><br>ODB:<br>1 prescription for a cholinesterase inhibitor |
| Frailty | Frailty:<br>Individuals were identified as having medical conditions associated with frailty based on health care encounters recorded in DAD, SDS, NACRS, and OHIP in the 2-years prior to index using the Johns Hopkins ACG® System Version 10.[10] |
| History of transient ischemic attack or acute ischemic stroke | This variable was included *a priori* as hypothesized to be directly related to COVID-19 infection risk.[3]<br><br>Transient Ischemic Attack:<br>DAD and NACRS were used to identify patients with a history of a transient ischemic attack, based on at least 1 hospitalization or ED visit with a diagnosis coded with one of the following codes:<br><br>ICD-9: 435, 3623<br>ICD-10: G450, G451, G452, G453, G458, G459, H340<br><br>Acute Ischemic Stroke:<br>DAD was used to identify patients with a history of acute ischemic stroke, based on at least 1 hospitalization with a main diagnosis coded with one of the following codes:<br><br>ICD-9: 434, 436; ICD-10: I63, I64, H34.1 |
| Influenza vaccine received, 2019-2020 season or 2020-2021 season | An OHIP billing with any of the following fee codes from October 1, 2019 to September 30, 2020, or October 1, 2020 up to 14 days before the index date: G590, G591, G592, Q130, Q590, Q690, Q691; |

| Variable | Definition |
|---|---|
| | or, an ODB billing with any of the following Drug Identification Numbers (DINs)/Product Identification Number (PINs) from October 1, 2019 up to September 30, 2020: 02420643, 02420783, 02432730, 02473283, or October 1, 2020 up to 14 days before the index date: 02420643, 02420783, 02432730, 02445646, 02494248, 09857645, 09857646 |
| Dissemination area (DA) | A dissemination area (DA) is the smallest standard geographic area for which all census data are disseminated. A DA generally comprises approximately 400-700 people, but in densely populated cities may contain several thousand people. DAs cover all the territory of Canada.[23]<br>We assigned subjects to a DA using postal code, as recorded in the Registered Persons Database. |
| Household income quintile | Calculated at the DA level using 2016 Census data[24] by multiplying the median income (before-tax) by the number of households and dividing by the sum of single-person equivalent to obtain income per single person equivalent. For DAs where median income was unavailable, neighbouring DAs were used to estimate income per single person equivalent. DA-based income quintiles were constructed separately for each census metropolitan area or census agglomeration (one or more adjacent municipalities integrated via commuting flows). DAs within each such area were ranked from the lowest average income per single-person equivalent to the highest, and DAs were assigned to five groups, such that each group contained approximately one-fifth the total in-scope population of each area. |
| Household density quintile (persons per dwelling) | Average number of persons in private households, calculated at the DA level using the 2016 Census data.[25] DAs across the province were ranked by average number of persons per household into 5 categories (quintiles), such that each group contained approximately one-fifth of the DAs. |
| Essential work quintile (proportion working in non-health essential services) | Calculated at the DA level, using 2016 Census data.[26] For each DA, we calculated the number of individuals ≥15 years old that were working in one of the following Census-defined work categories: Sales and service occupations; trades, transport and equipment operators and related occupations; natural resources, agriculture and related production occupations; and occupations in manufacturing and utilities.<br><br>DAs across the province were then ranked by these percentages into quintiles, with the lowest 1/5 of DAs comprising the first quintile, and so on. |
| Visible minority quintile (proportion self-reporting as visible minorities) | Calculated at the DA level, using 2016 Census data.[26] An individual was marked as "self-identify as a visible minority" if they reported being one or more of the following (wording from the 2016 Census): "South Asian (e.g., East Indian, Pakistani, Sri Lankan, etc.), Chinese, Black, Filipino, Latin American, Arab, Southeast Asian (e.g., Vietnamese, Cambodian, Laotian, Thai, etc.), West Asian (e.g., Iranian, Afghan, etc.), Korean, Japanese, or Other—specify".<br><br>DAs across the province were then ranked by these percentages into quintiles, with the lowest 1/5 of DAs comprising the first quintile, and so on. |
